# Advancing molecular, phenotypic and mechanistic insights of *FGF14* pathogenic expansions (SCA27B)

**DOI:** 10.1101/2024.01.15.23300194

**Authors:** Lars Mohren, Friedrich Erdlenbruch, Elsa Leitão, Fabian Kilpert, G. Sebastian Hönes, Sabine Kaya, Christopher Schröder, Andreas Thieme, Marc Sturm, Joohyun Park, Agatha Schlüter, Montserrat Ruiz, Moisés Morales de la Prida, Carlos Casasnovas, Kerstin Becker, Ulla Roggenbuck, Sonali Pechlivanis, Frank J. Kaiser, Matthis Synofzik, Thomas Wirth, Mathieu Anheim, Tobias B. Haack, Paul J. Lockhart, Karl-Heinz Jöckel, Aurora Pujol, Stephan Klebe, Dagmar Timmann, Christel Depienne

**Author notes:** Correspondence to: Christel Depienne, Institut für Humangenetik, Universitätsklinikum Essen, Virchowstraße 171, 45147 Essen, Germany. These authors contributed equally to this work (Lars Mohren, Friedrich Erdlenbruch, Elsa Leitão, Fabian Kilpert).

## Abstract

Repeat expansions in the *FGF14* gene have recently been identified as a frequent cause of autosomal dominant late-onset cerebellar ataxia (SCA27B). The threshold for pathogenicity was estimated to range from 250 (incomplete penetrance) to 300 AAG repeats (full penetrance) based on expansion sizes observed in patients and controls. However, the full sequences of pathogenic and non-pathogenic alleles remain largely unknown.

In this study, we used STRling and ExpansionHunter/STRipy to detect short tandem repeat expansions in short-read genome data of 80 patients with unsolved neurological disorders, including 48 patients from 39 unrelated families with cerebellar ataxia. Significant outlier values indicating a possible *FGF14* repeat expansion were detected in 49% of families with ataxia. We used long-range PCR and repeat-primed PCR to confirm repeat motifs and number, and to further screen for *FGF14* repeat expansions in 106 additional patients. The distribution of *FGF14* alleles was also analyzed in 802 control individuals. Long-range PCR amplicons from affected and control subjects with at least one expanded allele were sequenced by nanopore sequencing.

Altogether, we report a total of 38 individuals from 31 families with a pure AAG expansion ≥250 repeats in *FGF14* (range: 254-937) out of 134 families with unsolved ataxia (23%). Four families had biallelic expansions. One individual showed mosaicism of the expansion, with two distinct large alleles detected. The overall distribution of *FGF14* alleles significantly differs in patients and control subjects with pure expansions from 180 repeats being enriched in patients while AAGGAG and interrupted alleles were more frequent in controls. Expanded and non-expanded *FGF14* alleles were associated with different 5’-flanking regions correlating with repeat stability. Episodic symptoms of ataxia and downbeat nystagmus were frequent in patients with SCA27B independently of the repeat number. A family with a novel nonsense variant (NM_175929.3: c.239T>G; p.Leu80*; SCA27A) exhibited similar symptoms but earlier age at onset than patients with pathogenic expansions (SCA27B).

In conclusion, this study reveals the complete sequence of pathogenic and non-pathogenic *FGF14* expansions. We suggest that pure AAG expansions are pathogenic from a lower threshold than previously reported, comprised between 180 and 220 pure AAG repeats, while full penetrance would be above 320-335 repeats. Interrupted expansions and expansions composed of other hexanucleotide repeats motifs are non-pathogenic. Diagnostic tests should thus include a complete sequencing of the expansion in addition to repeat number assessment. Finally, we provide mechanistic insights into how pure AAG repeat expansions could lead to adult-onset cerebellar ataxia.

## Introduction

Spinocerebellar ataxias (SCAs) are a group of autosomal dominant, slowly progressive disorders characterized by impaired coordination and imbalance resulting in ataxia of stance and gait, limb ataxia, dysarthria and oculomotor signs^1^. Cognitive impairment, tremor, rigidity, bradykinesia, dystonia, spasticity, and polyneuropathy are frequently associated features. So far, at least 40 different SCA subtypes, classified according to their underlying locus/genetic cause, have been reported^2^. This list includes repeat expansions of CAG trinucleotides encoding polyglutamine (PolyQ) repeats (SCA1, 2, 3, 6, 7, 12, 17) as well as noncoding repeat expansions of penta- or hexanucleotides (SCA10, 12, 31, 36, 37). Furthermore, point variants have been described in at least 28 distinct genes^1,2^. Among those, rare point variants or microdeletions leading to the heterozygous loss-of-function of *FGF14* had previously been reported as SCA27 (renamed SCA27A)^3–5^.

More recently, heterozygous noncoding AAG/CTT repeat expansions in *FGF14* have been identified as a frequent cause of late-onset cerebellar ataxia (SCA27B) in Canada, Australia, and Europe^6–8^. *FGF14* encodes the fibroblast growth factor 14, a gene expressing at least eight different isoforms, according to the Ensembl database. The Matched Annotation from the NCBI and EMBL-EBI (MANE) isoform (ENST00000376143.5; NM_004115.4), also called transcript 1, encodes a 247-amino acid (27 kDa) protein that is considered as the canonical sequence in Uniprot. However, this transcript is poorly expressed in all tissues according to the Genotype-Tissue Expression (GTEx) database. The most abundant transcript (transcript 2; ENST00000376131.9; NM_175929.3) is brain-specific, with the highest expression in the cerebellum, and encodes a 252-amino acid (28 kDa) protein. The two protein isoforms differ in their N-terminus as a result of distinct transcription start sites, leading to the inclusion of different first exons: the first 64 amino acids of isoform 1 (FGF14-1a) are substituted with an alternative sequence of 69 amino acids in isoform 2 (FGF14-1b)^9^. The N-terminus of isoform 1 contains a nuclear localization signal and is predicted to localize in the nucleus. In contrast, isoform 2 localizes at the axon initial segment of cerebellar Purkinje neurons^10^, where it regulates the activity of voltage-gated sodium (Nav1.2/*SCN1A* and Nav1.6/*SCN8A*)^9^ and potassium (Kv7.2/*KCNQ2* and Kv7.3/*KCNQ3*) channels^11^. The repeat expansion responsible for SCA27B is located in intron 1 of isoform 2. The expansion is usually described as an AAG (or GAA) repeat expansion according to its genomic context, but as the gene is on the reverse strand, the expansion consists of CTT repeats in the gene context. The expansion occurs at a short tandem repeat highly polymorphic in humans and is associated with a probable founder effect in Canada, with an incidence up to 60% of families in this population^6^. The comparison of expansion sizes in patient and control populations has suggested that expansions above 300 AAG repeats are pathogenic with full penetrance while expansions between 250 and 299 repeats are associated with incomplete penetrance^6,7^. The consequence of the expansion is a reduction of *FGF14* expression from the expanded allele, likely leading to the heterozygous loss-of-function (haploinsufficiency) of isoform 2 in the cerebellum^6^.

In this study, we identify *FGF14* repeat expansions as the most frequently missed genetic cause of cerebellar ataxia in a cohort of patients with neurological disorders who had previous inconclusive exome or genome analyses. This finding led us to analyze *FGF14* alleles in a total of 988 individuals, including 154 patients from 134 independent families with cerebellar ataxia of unknown cause, 32 patients with other neurological disorders, and 802 control subjects, using a combination of long-range PCR, fluorescent gene fragment analysis and targeted nanopore sequencing. Our results emphasize the importance of sequencing both the repeat expansion and its flanking region for an accurate clinical interpretation. We suggest that thresholds established for defining pathogenic and likely pathogenic repeat thresholds should be reassessed. Moreover, we show that patients with 250-299 repeats and patients with ≥300 repeats have indistinguishable clinical features. Additionally, we provide mechanistic insights into how pure AAG repeat expansions lead to adult-onset cerebellar ataxia while expansions of other motifs or interrupted expansions are non-pathogenic.

## Materials and methods

### Patients & subjects

Patient inclusion was part of the project “Identification of tandem repeat EXPAnsions in unsolved Neurological Disorders” (EXPAND). The study has received the approval of the ethics committee of University Hospital Essen (21-10155-BO). All patient and subjects’ consent was obtained according to the Declaration of Helsinki. At the time of the study, 76 patients with neurological disorders remaining without any identified genetic cause after an exome or a genome analysis were recruited from the Department of Neurology, University Hospital Essen, and had their genome sequenced. Among these, 44 patients from 35 independent families had spinocerebellar ataxia as a main clinical feature. Additionally, data of four families with cerebellar ataxia from Spain were included in the analysis. The *FGF14* repeat expansion was screened for in an independent cohort of 95 index cases with cerebellar ataxia without available genome data and 11 additional affected family members. One patient had a point variant in *FGF14* (NM_175929.3: c.239T>G; p.Leu80*) identified by routine exome sequencing. This study also included 802 control subjects: 30 anonymous blood donors and 772 participants from the Heinz-Nixdorf Recall (HNR) Study^12^. Patient and family identification numbers (IDs) were used for pseudonymization and were not known to anyone outside the research group.

### Genome sequencing & expansion calling

Short-read genome sequencing was performed at the Cologne Genome Center (Cologne, Germany) for 51 patients and as part of routine diagnosis at the Institute for Medical Genetics and Applied Genomics (University of Tübingen, Germany) for 25 patients. In all patients, Fastq data were mapped to the hg38 reference genome using an in-house pipeline (Supplementary Methods). We used STRling^13^ (version 0.5.2), ExpansionHunter^14^ v5.0 and STRipy^15^ for repeat expansion detection.

### Long-range PCR & Repeat-primed PCR

*FGF14* Repeat expansions were amplified by long-range PCR (LR-PCR) and repeat-primed PCR (RP-PCR) using protocols adapted from Rafehi *et al*^7^ (Supplementary Methods). Notably, we reduced the number of cycles of the LR-PCR to limit an enhanced amplification of smaller alleles. Amplification was performed with a FAM-labelled forward primer for gene fragment analysis on ABI 3130xl DNA Analyzer (Applied Biosystems, Waltham, MA). The sizes of *FGF14* alleles below 700-1200 bp and not sequenced by nanopore sequencing were analyzed using GeneMarker (SoftGenetics LLC, PA).

### Nanopore sequencing

LR-PCR products were sequenced for 123 individuals: 100 subjects (53 patients with cerebellar ataxia and 47 control subjects) with an allele ≥700 bp, and 23 samples (17 controls and 6 patients) with an allele between 487 and 685 bp (due to discrepancies between values estimated on agarose gel and fragment analysis). LR-PCR amplification was performed without FAM-labelled primer in a total volume of 75 µl. Amplicons were purified using the DNA Clean & Concentrator (Zymo Research, Irvine, CA). We used the SQK-LSK109 ligation-based sequencing kit (Oxford Nanopore Technology, Oxford, UK) and the native barcoding protocol (EXP-NBD196) to prepare the libraries and multiplex samples. Basecalling was performed using a command line in-house Snakemake^16^ workflow available at GitHub (Supplementary Methods). Reads were visually inspected with GeneiousPrime®2019 (Biomatters Ltd.) for identification of the variable 5’ flanking region, pre-repeat, main motif, additional repeat motif, and interruptions. Differences between alleles were used to separate reads into a1 (smaller), a2 (larger) and a3 (mosaic cases) alleles.

### Circular dichroism (CD) spectroscopy

We used CD spectroscopy to investigate the structure of AAG and AAGGAG repeats and complementary counterparts (Supplementary Methods). CD spectra were measured over a spectral range of 200-340 nm on a Jasco J-710 CD spectropolarimeter coupled to a Jasco PFD-3505 Peltier temperature controller. Measurements were carried out at 20 °C in a 1 mm path length quartz cuvette with a scanning speed of 200 nm/min, response time of 2 s, a bandwidth of 1 nm, and an accumulation of four spectra.

## Results

### Identification of *FGF14* expansions from genome data

We used STRling to identify STR expansions in short-read genome data of 80 patients with neurological disorders (76 from Germany and 4 from Spain), including 48 patients with cerebellar ataxia from 39 independent families (Fig. 1). This analysis detected outlier values associated with significant q values indicating possible *FGF14* AAG repeat expansions (chr13(hg38): 102,161,567-102,161,726) in 23 of the 48 patients with ataxia (19/39 families; 49%). Significant values were detected for both individuals of four families for which genome data were available. In contrast, only three of the 32 individuals with other neurological disorders (3/31 families; 10%) showed significant outlier values. In addition, STRling detected an expansion of another motif (AAGGAG) at the same locus in one family from Spain.

**Figure 1.**
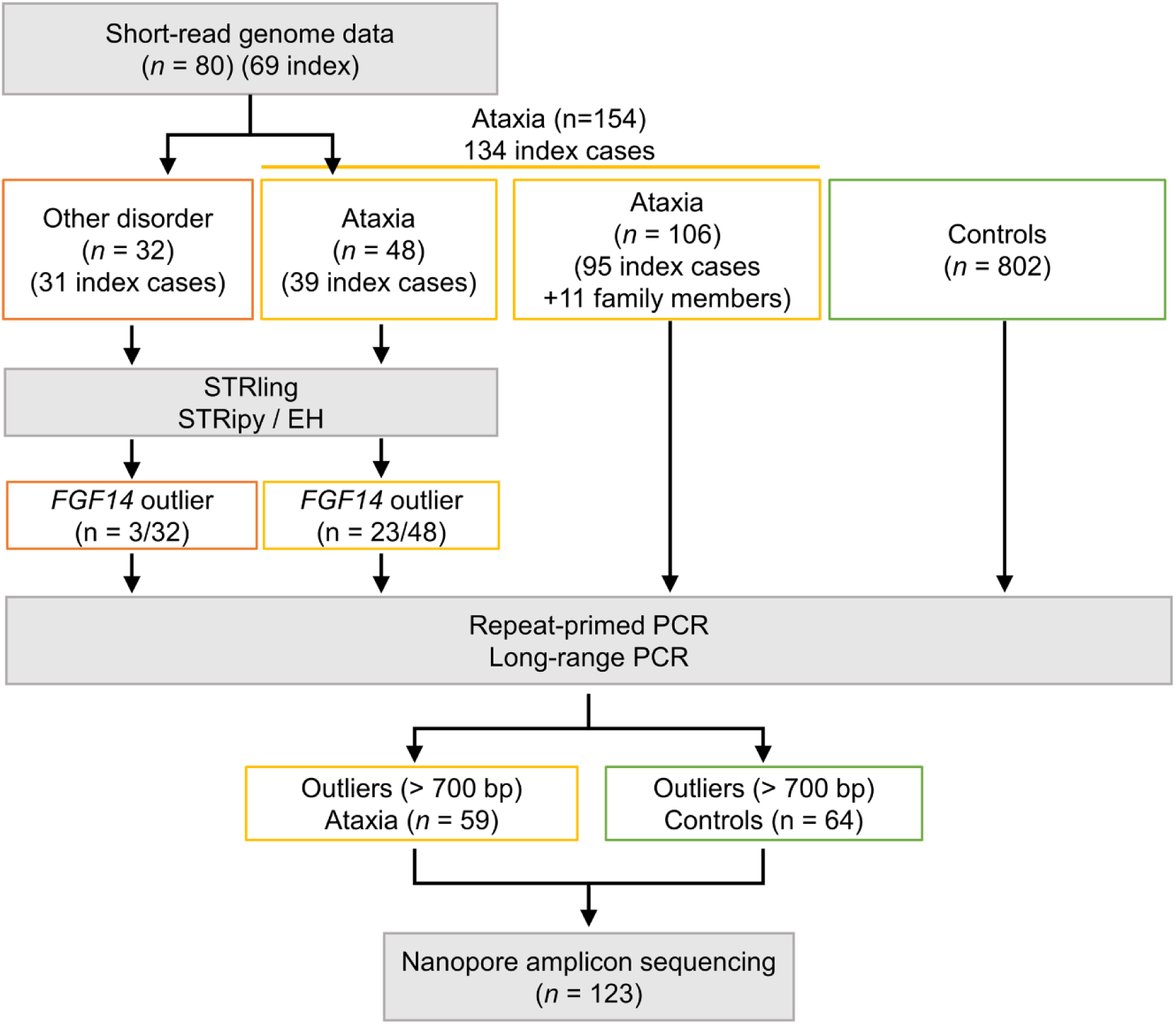
Flowchart illustrating the design of the study. This figure visually represents the sequential steps, methods used, and distribution of participants at each stage.

We set up LR-PCR and RP-PCR assays to validate these results. The existence of at least one large *FGF14* AAG allele (PCR product ≥700 bp; triplet repeat number ≥180) was confirmed for 18 of the 26 individuals, all with cerebellar ataxia. The eight remaining individuals, including the three with another neurological disorder, had a larger allele below 650 bp (i.e. ≤160 repeats) and were therefore considered negative for *FGF14* expansion/SCA27B. Of note, we also confirmed that all patients that had no outlier values detected by STRling only had small *FGF14* alleles (Supplementary Fig. 1).

### Screening of expansions by LR-PCR and RP-PCR

We then used the LR-PCR and RP-PCR assays to analyze 106 additional patients (95 new index cases and 11 family members) with cerebellar ataxia (Fig. 1, Fig. 2A and Supplementary Fig. 2; information on pedigrees on request). Twenty-six of the 95 index cases (27%) and five family members had at least one AAG allele ≥700 bp. Taken together with the expansions detected from genome data, 49 individuals from 40 families out of 134 independent families analyzed (30%) had an expanded AAG allele (≥180 repeats) estimated from fragment size or gel analysis. The AAG expansions segregated with the disorder in all families including at least two affected individuals available for genetic analysis. Four patients from three families had an expansion of the AAGGAG motif (Supplementary Fig. 2B). The AAGGAG expansion segregated in two affected individuals from a Spanish family but was present in only the index case but not the affected parent of a German family and was further considered as non-pathogenic, as already reported^6,7,17,18^.

**Figure 2.**
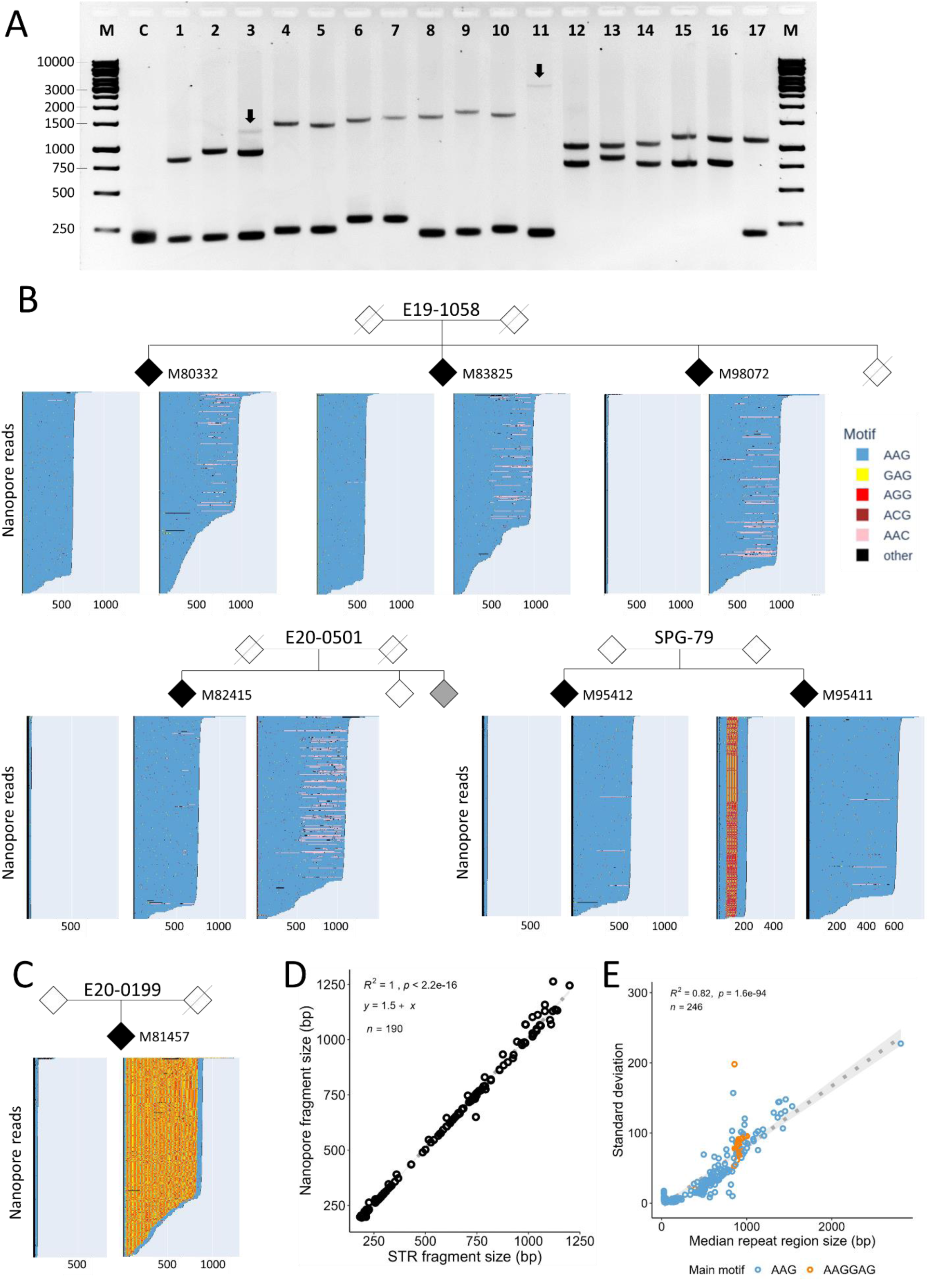
Analysis of *FGF14* repeat expansions in affected subjects using LR-PCR and nanopore sequencing. **A)** Gel electrophoresis of selected LR-PCR products spanning the *FGF14* (AAG) STR locus. From left to right, C: negative control; lanes 1-2: examples of *FGF14* expansions between 220 and 299 repeats ; lane 3: individual showing one small and two large alleles (somatic mosaicism of the largest allele); lanes 4-11: examples of *FGF14* expansions above 300 repeats (patient in line 11 with 937 repeats); lanes 12-16: individuals with biallelic expansions (15-M83825, 16-M80332); lane 17 corresponds to the sibling of individuals 15 and 16 who both have biallelic expansions (family E19-1058, M98072); the third sibling has a heterozygous expansion of 325 repeats. **B)** Schematic representation of nanopore reads sequenced for selected family members. *FGF14* alleles have been separated based on their flanking regions (see methods). 300 randomly chosen reads are displayed in each graph. AAG repeats appear in blue. GAG, AGG, and AAC repeats appear in yellow, red and light pink respectively. Other sequences appear in black. The panel above shows the segregation of the *FGF14* alleles in family E19-1058 comprising three affected siblings, two with biallelic expansions and one with a single expanded allele. Lower-left panel: individual M82415 (E20-0501) showed reads that could be partitioned into three different alleles: one small allele and two large alleles (somatic mosaicism). Lower-right panels: family SPG-79 including two affected family members with intermediate FGF14 alleles (203 and 207 repeats). Note interruptions in the middle of the same allele of individual M95411. **C)** Nanopore reads from an individual with a small and an expanded AAGGAG allele. **D)** Correlation between the median number of repeats detected by nanopore sequencing and expansion size estimated from fragment size analysis. Due to the very high correlation between expansion sizes detected by nanopore sequencing and fragment size analysis, we could confidently use the values calculated by nanopore sequencing to fill in missing allele values > 700-1200 bp (i.e. alleles too large to be analyzed by fragment size analysis). **E)** Standard deviation in allele size calculated from nanopore reads showing that somatic instability is positively correlated with expansion size. Patient/family IDs are not known to anyone outside the research group.

### Sequence of expansions ≥250 repeats

Since both LR-PCR and RP-PCR failed to provide the precise count of AAG repeats at the *FGF14* locus, we developed a multiplex nanopore sequencing assay of the LR-PCR amplicons. In parallel, we also analyzed and compared the distribution of *FGF14* alleles in a control cohort composed of 802 subjects. In total, we sequenced *FGF14* alleles in 59 patients with cerebellar ataxia and 64 control individuals (Fig. 1; Fig. 2B-C). We observed a strong correlation between the allele sizes determined through fragment analysis or gel electrophoresis and the median number of repeats calculated from the nanopore data (Fig. 2D). Taken together, fragment analysis and nanopore sequencing of LR-PCR amplicons allowed us to have a complete overview of the size of both alleles present in patients and control subjects.

Overall, thirty-eight patients from 31 families (31/134; 23%) had at least one allele composed of pure AAG repeats exceeding the current established threshold for pathogenicity (250 repeats) and were considered as having SCA27B. The median number of repeats calculated from nanopore reads ranged from 254 to 937 AAG repeats (Supplementary Table 1). Twenty-six patients had a repeat number above 300 repeats, whereas 12 patients had a repeat number between 250 and 299. Nevertheless, we noted that the thresholds of 250 and 300 repeats appeared somewhat arbitrary. For instance, in one family, two siblings with similar symptoms and age at onset (AAO) had median repeat numbers of 401 and 281 repeats, respectively. Similarly, in another family, the index case had 258 repeats and the affected parent had 224 repeats (information on pedigrees on request).

Five patients from four independent families showed biallelic repeat expansions (Fig. 2A-B). In three families, the largest allele was below 300 repeats and the lowest allele below 250 repeats: 222/278, 196/284, 165/272. The fourth family included three affected siblings; two siblings had 204/311 and 196/319 repeats whereas the third sibling only had one large pathogenic allele (325 repeats). One patient repeatedly exhibited two large alleles, one with ≥300 repeats and another between 250 and 300 repeats, in addition to a small allele, suggesting somatic variability of the expanded allele (Fig. 2A-B). More generally, a high degree of somatic mosaicism around the mean value was detected for all individuals with repeat expansions, as highlighted by the positive correlation between the standard deviation and the allele size (Fig. 2E).

### Intermediate *FGF14* alleles

Eleven patients had a *FGF14* AAG repeat expansion between 180 and 249 repeats (information on pedigrees on request). Using the current threshold of 250 repeats, these patients are considered negative for SCA27B. As the parent of the index patient with 258 repeats was affected with a median value of 224 repeats and we had genome data for both members of this family, we investigated whether there might be another genetic cause for her ataxia, but we could not identify any variant associated with ataxia other than the *FGF14* expansion. We thus investigated the possibility that alleles below 250 might also be associated with disease. Most pedigrees of patients with intermediate alleles were uninformative but we observed that the expansion segregated in a Spanish family including two affected individuals respectively (information on pedigree on request). One patient had a variant of unknown significance in *PUM1* (NM_001020658.2: c.2180T>C; p.(Ile727Thr)) in addition to 236 AAG repeats in *FGF14*. The two other families had no variant detected in genes associated with ataxia. Among the patients without genome data, one isolated patient had a pathogenic SCA6 expansion (21 repeats) in addition to 209 repeats in *FGF14*.

### Comparison of *FGF14* alleles in patients and controls

The 802 control subjects showed an overall different distribution of *FGF14* alleles compared to patients with ataxia (Fig. 3A-B), especially when considering only the larger allele (Supplementary Fig. 4B; Mann-Whitney, p=0.0002). Large alleles composed of pure AAG ≥180 repeats were enriched in patients with ataxia with alleles ≥250 repeats showing a more significant enrichment than intermediate alleles (Fig. 3C-D). Conversely, AAGGAG expansions were more frequent in controls (Fisher’s test: *p*=0.02, OR 3.8; Fig. 3A). We observed true AAG interruptions (disrupting repeats in the middle) in smaller alleles only, while interruptions limited to 3’ or 5’ sides of the repeats were equally frequent in both alleles (Fisher’s test: *p*=0.47) but more frequent in control subjects than patients with ataxia (Fisher’s test: *p*=0.01, OR 4.2; Fig. 3; Supplementary Fig. 3). Out of 21 control subjects who had at least one allele above 250 repeats, 13 were composed of the non-pathogenic AAGGAG repeat motif (Supplementary Fig. 3; Supplementary Fig. 4). Eight control individuals had repeat expansions ≥250 AAG repeats and only two, aged 46-50 years old and 66-70 years old at the time of sampling, had a pure AAG repeat expansion above 300 repeats (313 and 319 repeats respectively; Supplementary Table 1).

**Figure 3.**
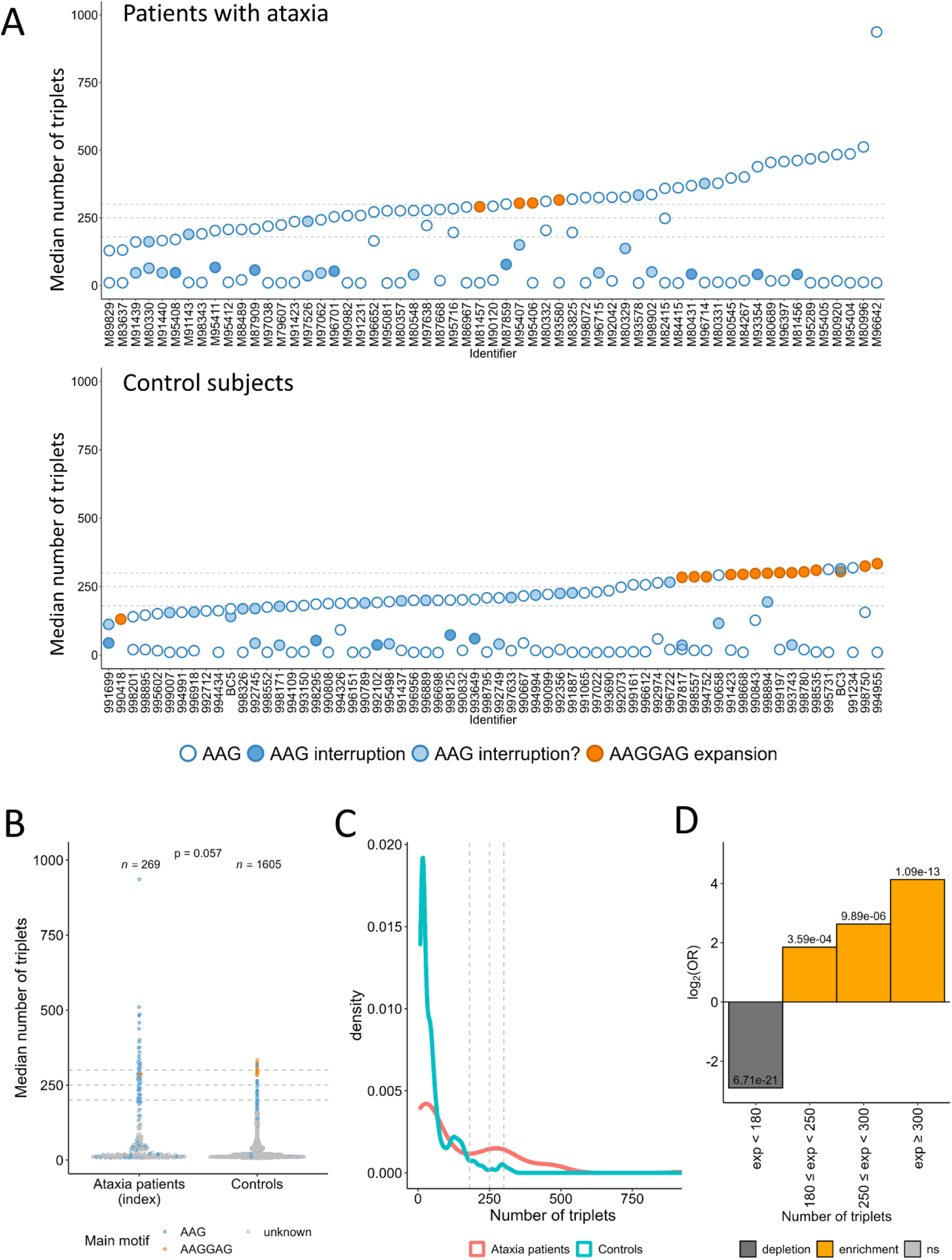
Distribution of *FGF14* alleles in patients with cerebellar ataxia and control subjects. **A)** Median number of triplets of both alleles for the 59 patients with ataxia and 64 control individuals sequenced by nanopore sequencing. Pure AAG alleles are depicted as blue dots with a white fill. AAGGAG alleles appear in orange. Alleles with interruptions are depicted as blue dots with a dark blue fill. Alleles with interruptions limited to the 5’ or 3’ of the expansion are depicted as blue dots with a light blue fill. **B)** Comparison of median allele sizes (including all alleles of each individual) in patients with cerebellar ataxia and control subjects. **C)** Density plot showing the different distributions of the number of triplets in the larger allele for ataxia patients and controls. **D)** Log odds ratio according to repeat numbers in all alleles (ataxia patients versus controls) showing a significant enrichment of alleles > 180 repeats in patients with cerebellar ataxia. Figures similar to B) and D) but taking only the large alleles into account, appear in Supplementary Figure 4. Patient IDs are not known to anyone outside the research group.

### Variability of the flanking region

Interestingly, the 5’ region flanking the repeats (3’ region in the context of the gene) was highly variable and drastically differed in expanded versus non-expanded alleles (Fig. 4A). We observed seven different sequences following a constant CTTTCT motif (chr13:102,161,558-102,161,563) upstream of the repeats. These 5’-flanking sequences were either directly followed by AAG repeats, or preceded by short AG-rich sequences (e.g., AAGAAAGAG or AAGAG) that we considered as ‘pre-repeat’ (Fig. 4B; Supplementary Fig. 5A-B). Pre-repeats were more frequently observed in larger (a2) alleles (Fisher’s test: *p*=3.227×10^-8^, OR 7.5; Fig. 4C; Supplementary Fig. 5C-D). The variable GTG sequence (present in the hg38 reference genome) was the most frequently associated with *FGF14* expansions although the range of repeats observed was highly variable (range: 36-512). GG and GGG sequences were only detected in expanded alleles (326-937 repeats). Conversely, the GTTAGTCATAGTACCCC was strikingly associated with small alleles (9-21 triplet repeats) only. Interestingly, one nearly identical sequence, differing only in the final two nucleotides (GTTAGTCATAGTACCAG), was associated with 203/207 repeats in both affected individuals of family SPG-79. This suggests that the four consecutive cytosines at the end of the sequence play a key role in preserving the stability of the adjacent repeats.

**Figure 4.**
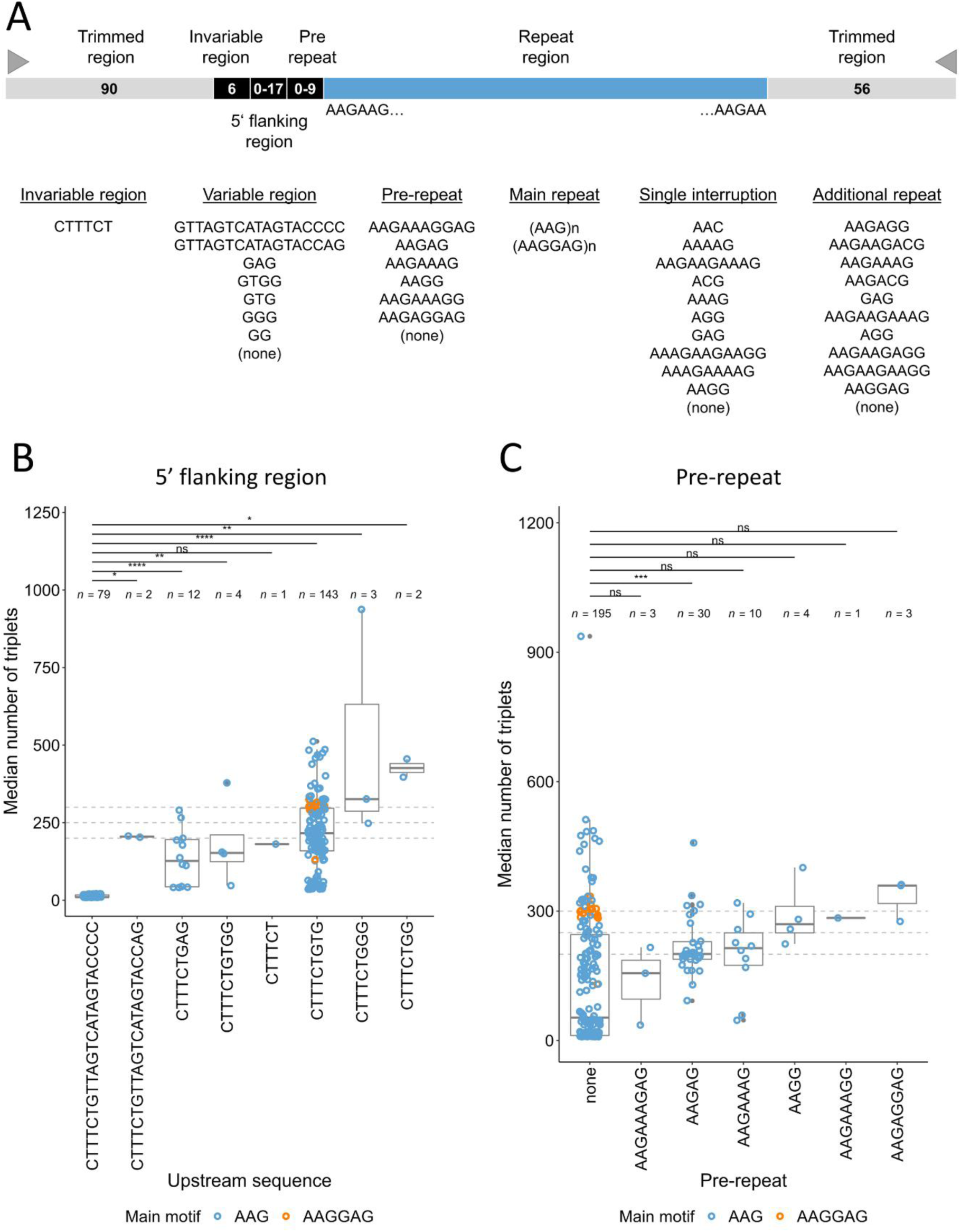
Effect of 5’ flanking regions on repeat instability. **A)** Schematic representation of the different parts composing *FGF14* repeat expansions. An invariable CTTTCT motif is usually followed by a variable 5’ region. A pre-repeat can be present in some individuals before the repeats. Some alleles are interrupted by one or several other motifs called interruptions. **B)** Median number of triplets for each allele depending on the flanking region sequence. GTTAGTCATAGTACCCC is present in small alleles (≤21 repeats) only. Other sequences show higher number of repeats, suggesting higher instability of these associations. **C)** Median number of triplets for each allele depending on the pre-repeat motif. Graphs displayed in panels B) and C) include both patients with ataxia and controls. Graphs presenting data for patients with ataxia and controls separately appear in Supplementary Figure 5.

### Clinical features associated with *FGF14* expansions

We divided patients with cerebellar ataxia into four distinct groups for clinical comparison: 1) patients with a median number of AAG repeats ≥300; 2) patients with 250-299 median repeats; 3) patients with an intermediate allele (180-249); and 4) patients negative for SCA27B (detailed clinical features of individual patients on request). Furthermore, we included the clinical data of a German family (affected parent-child pair) with a novel pathogenic nonsense variant in *FGF14* (SCA27A; NM_175929.3:c.239T>G; p.(Leu80*); NM_004115.4 (MANE):c.224T>G, p.(Leu75*);) identified by routine exome sequencing (Fig. 7A-B).

Overall, most patients with *FGF14* expansion ≥250 repeats (29/38; 76%) had a highly recognizable phenotype, characterized by the association of slowly progressive cerebellar signs accompanied by episodic symptoms of ataxia and/or downbeat nystagmus (DBN) that often present as first symptoms. In one patient, symptoms were limited to episodic ataxia without symptoms in between episodes. In nine patients, cerebellar symptoms were present without episodic features or DBN. In six patients, the phenotype was associated with additional signs or an overall different course of the disease. One patient (#5 in Fig. 6F-G) had episodic choreatiform movements, increased muscle tone of the lower limbs, in addition to cerebellar signs and DBN. Patient #1 had spasticity in the legs. Patient #4, with biallelic expansion, cerebellar ataxia was accompanied by an anxious/depressive disorder. The patient also had a stroke after first presentation from which she completely recovered (without significant change on SARA scores). SARA scores, however, may have been biased by anxiety-induced exacerbation of the patient’s balance problems. Patient #6 had a slowly progressive cerebellar syndrome with mild autonomic symptoms over 20 years. Later on, the patient developed idiopathic Parkinson’s disease and dementia. Two patients with high SARA scores showed moderate to severe cerebellar atrophy. In addition, one (#2) of these two patients had surgery for epidermoid tumor of the basal cisterns and seizures as well as multiple meningiomas and developed leukoencephalopathy and dementia in later age. The other patient (#3) had a history of pulmonary sarcoidosis with muscular involvement (with no muscular involvement at examination). Detailed case reports of these individuals are available on request.

Patients with more than 300 repeats and patients with 250-299 repeats exhibited comparable clinical characteristics that slightly differed from *FGF14*-negative patients with ataxia. For example, we observed a significantly higher occurrence of early cerebellar oculomotors signs (95%; 92% and 96%, respectively). In particular, DBN was present in 47% (50% and 46%) at the first examination, compared to only 3% in patients negative for *FGF14* (Fig. 5A). Patients with SCA27B had a lower occurrence of dysarthria on the first examination (24%; 25% and 23%, respectively) compared to 62% in the *FGF14* negative group; Fig. 5A). There was less cognitive impairment in patients with *FGF14* expansions compared to the *FGF14* negative group either at the first or last examinations (71% of negative patients had cognitive impairment versus 38% in the *FGF14*-positive group; Fig. 5B). These statistical differences remained even when removing the six patients behaving as clinical outliers.

**Figure 5.**
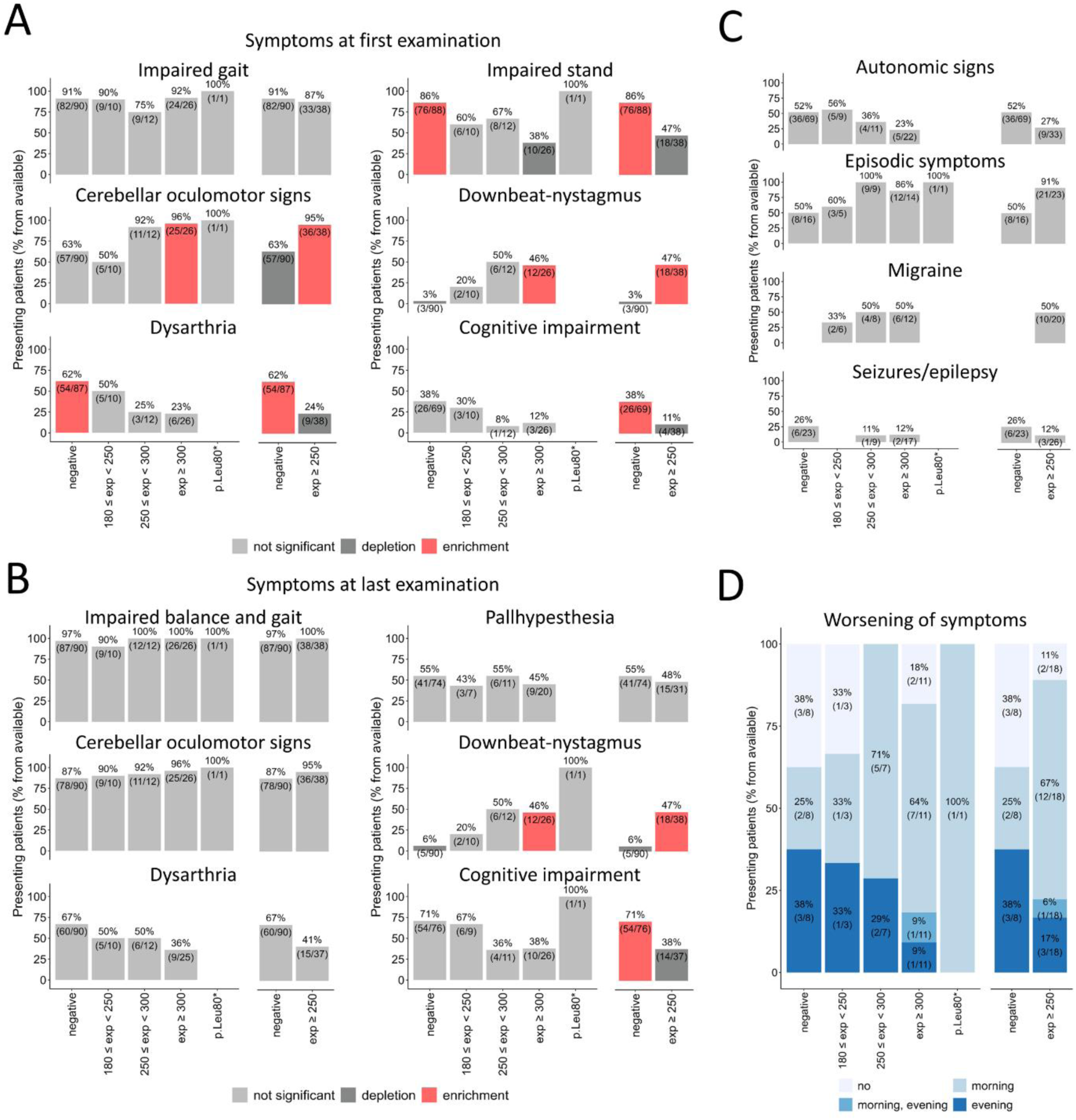
Clinical characteristics of individuals with *FGF14* pathogenic expansions and point mutations. **A)** Bar graphs showing the percentage of patients presenting impaired gait, impaired stand, cerebellar oculomotor signs, downbeat-nystagmus, dysarthria and cognitive impairment at first examination. **B)** Bar graphs showing the percentage of patients presenting impaired gait and stand, cerebellar oculomotor signs, downbeat-nystagmus, dysarthria, pallhypersthesia and cognitive impairment at last examination. **C)** Bar graphs showing the percentage of patients presenting autonomic signs, episodic symptoms, migraine, and seizures/epilepsy. For all graphs presented in panels A, B) and C), categories showing significant enrichment appear in red, significant depletion in dark gray while non-significant differences appear in light grey (Fisher’s test). **D)** Bar graphs showing the percentage of patients presenting a worsening of symptoms on morning, evening, morning and evening or no worsening of symptoms depending on the day time.

The progression of SCA27B ataxia, as monitored by SARA and ICARS scores, was globally slow with a mean increase in SARA scores of 8.7 points over 30 years (Fig. 6F-G). When removing the six patients considered as ‘clinical outliers’, the progression was even slower (4.5 points over 30 years; Supplementary Fig. 6D-E). However, the variability observed was high in both SCA27B groups (250≤repeats< 300 and ≥300 repeats). In total, the linear regression model suggests that disease duration only accounts for 7.9% of the variance observed, indicating that other factors, whether genetic or non-genetic, exert a more substantial impact.

**Figure 6.**
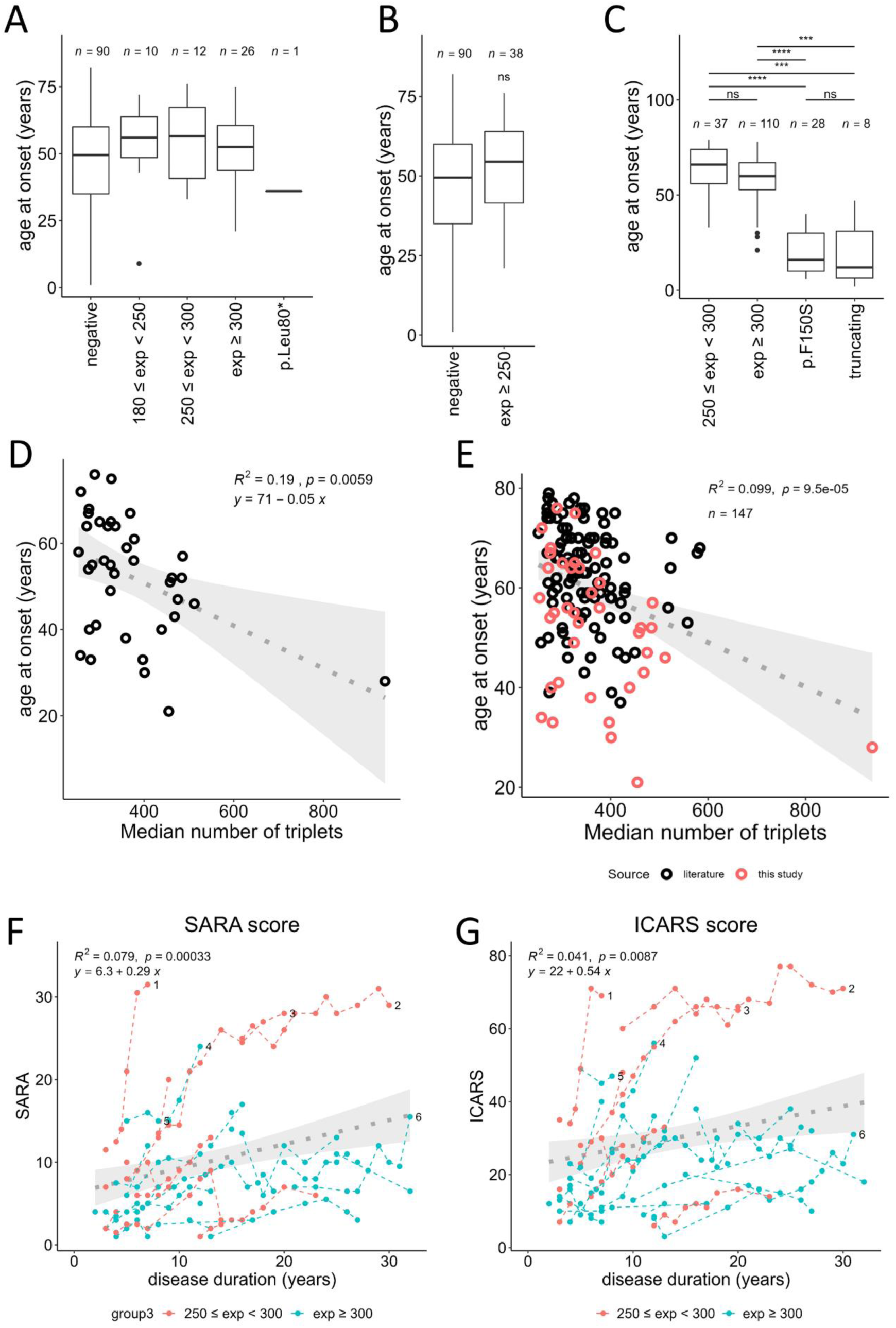
Analysis of the age at onset (AAO) and disease progression in patients with SCA27B/SCA27A. **A)** Comparison of the AAO in patients negative for SCA27A/SCA27B, patients with intermediate alleles (180-249), patients with *FGF14* expansions comprised between 250 and 299 repeats, patients with repeat size ≥300 repeats and the two patients with p.(Leu80*). **B)** Comparison of the age at onset in patients negative for SCA27A/SCA27B, and patients with SCA27B (repeat size ≥250 repeats). **C)** Meta-analysis comparing the age at onset in patients with *FGF14* expansions comprised between 250 and 299 repeats, patients with repeat size ≥300 repeats, patients with nonsense or frameshift variants in FGF14 or patients with p.Phe150Ser, showing that patients with pathogenic point variants (SCA27A) have an earlier age at onset than patients with repeat expansions (SCA27B). **D)** Correlation between the age at onset and *FGF14* AAG repeat number including only patients from this study. **E)** Correlation between the age at onset and *FGF14* AAG repeat number taking all patients from this study (red) and patients from previous studies (black) into account. **F)** SARA scores of patients with *FGF14* repeat expansions. **G)** ICARS scores of patients with *FGF14* repeat expansions. In both graphs shown in panels F) and G), patients with *FGF14* expansions comprised between 250 and 299 repeats appear in red while patients with ≥ 300 repeats appear in blue. Scores from the same patients at different time points are connected with dashed lines. Numbered last data points mark lines corresponding to atypical patients.

Interestingly, a large proportion of SCA27B patients reported a worsening of their symptoms on mornings (67%; 71% and 64% in *FGF14* positive groups compared to only 25% in the *FGF14* negative group; Fig. 5D). More than half of the patients who received 4-aminopyridine/fampridine reported an improvement of their symptoms (57%; 62% and 53%, respectively; Supplementary Fig. 6F-G). Treatment response to acetazolamide was also reported (29 %; 0% and 50%, respectively).

The patient harboring the pathogenic nonsense variant in *FGF14* (p.Leu80*) exhibited symptoms very similar to the groups of patients with ≥250 repeats. The patient had a slowly progressive cerebellar syndrome, episodic worsening of cerebellar symptoms and DBN. In addition, the patient also experienced episodic dystonia of the left hand. While the patient developed his first symptoms in adulthood (36-40 years), the child had tremors in childhood (1-5 years) and developed gait instability in adulthood (26-30 years).

Patients with intermediate repeat sizes (180-249 repeats) showed no significant differences from the groups of patients with ≥250 repeats, but this group appeared to be more similar to the negative group in terms of overall symptoms. One of the two patients with DBN was the parent of a patient with 258 repeats. Three other patients exhibited episodic features. Taken with the molecular findings, this suggests that some patients had an additional or different genetic cause and that the *FGF14* intermediate alleles would likely act as susceptibility factors in combination with other genetic or non-genetic factors.

### Correlations between repeat number and age at onset

The mean AAO in our cohort was 55.2 years (range 33-76; Fig. 6A) in patients with 250-299 repeats and 51.0 years (21-75 years) in patients with ≥300 repeats, whereas patients of the negative group had a mean AAO of 46.0 years (1-82 years). However, because of the high variation in all groups, the difference was not significant. Seven patients with an expansion presented with an earlier form of the disease, with an onset before 40 years old (21-38); all but two had ≥300 repeats. Accordingly, we observed a significant inverse correlation between the number of AAG repeats and the AAO (Fig. 6D; Supplementary Fig. 6A). Nevertheless, 81% of the variation is independent of the number of AAG repeats (*R*^2^=0.19). The high variability of the AAO for the same range of repeats was illustrated by a patient (258 repeats) who started to show symptoms in early adulthood (31-35 years) whereas another individual (259 repeats) only experienced the first symptoms in late adulthood (70-75 years).

We performed a meta-analysis of the correlation between the AAO and expansion size by pooling data of patients with *FGF14* expansions from our study and from four previous studies^7,19–21^. We also included data of patients with truncating variants^4,5,22,23^ or the F150S (F145S in MANE isoform 1) missense variant in *FGF14*^3,24^ in the comparison (information on age at onset on request). Altogether, we could confirm a significant inverse correlation between AAO and expansion size (Fig. 6E; supplementary Fig. 6B). The aggregated data also showed a tendency for patients with *FGF14* expansions between 250 and 299 repeats to be later affected on average (62.5 years; *n*=37) than patients with ≥300 repeats (59.1 years; *n*=110), but despite increased statistical power, the difference remains not significant due to the high variation in both groups (Fig. 6C). However, patients with expansions from both groups had a significantly later AAO compared with patients with truncating (19.4 years, range: 2-47) or F150S (20.5 years, range: 6-40) variants (Fig. 6C).

### Frequency of SCA27B in the German cohort

To evaluate the frequency of SCA27B compared to other dominantly inherited ataxia subtypes, we compared the number of patients who received a positive diagnosis in the ataxia outpatient clinic (Department of Neurology, University Hospital Essen). Forty patients had an expansion in *CACNA1A* (SCA6), 36 had SCA27B, 35 had an *ATXN3*/SCA3 expansion, 17 had an *ATXN1*/SCA1 expansions, 12 had a point variant in *CACNA1A* (Episodic Ataxia 2), seven had a variant in *PRKCG* (SCA14), six had an *ATXN2*/SCA2 and four had *ATXN8OS*/SCA8 expansions. Furthermore, eight had rarer forms (SCA28 and SCA49, two each; SCA7, SCA13, SCA15, SCA27A, one each). Altogether, this suggests that SCA27B is one of the most frequent SCA subtypes, accounting for approximately one-fifth of all diagnoses made in patients with cerebellar ataxia. This result independently confirms the high frequency of SCA27B diagnoses observed in another German cohort.^25^

### Secondary structures associated with *FGF14* expansions

We used CD spectroscopy to assess the potential of secondary structure formation of the different *FGF14* antisense repeat expansions AAG and AAGGAG as well as the complementary sense sequences CTT and CTCCTT on DNA and RNA. The AAG-DNA 25-mer formed an antiparallel homoduplex while CD spectroscopy of the AAGGAG-DNA oligo revealed formation of a parallel homoduplex^26–29^(Fig. 7C). The RNA counterparts CUU and CUCCUU did not obtain any secondary structure under the tested conditions, confirmed by a single positive band at 270 nm^30^ (Fig. 7C). Interestingly, the non-pathogenic AAGGAG-RNA oligo folded into a parallel guanine-quadruplex (G4) with a positive band around 260 nm, a negative band around 240 nm and positive values around 210 nm. Presence of a G4 was further confirmed by a G4-specific decrease in the stability detected, shifting from a parallel G4 structure (with 100 mM K) to a hairpin structure (with 100 mM Li)^31,32^ (Fig. 7C). Of note, for the pathogenic AAG-RNA repeat we detected a CD spectrum related to an A-form RNA structure, with a negative peak around 210 nm that reflects intra-strand interaction of RNA duplexes^33^.

**Figure 7.**
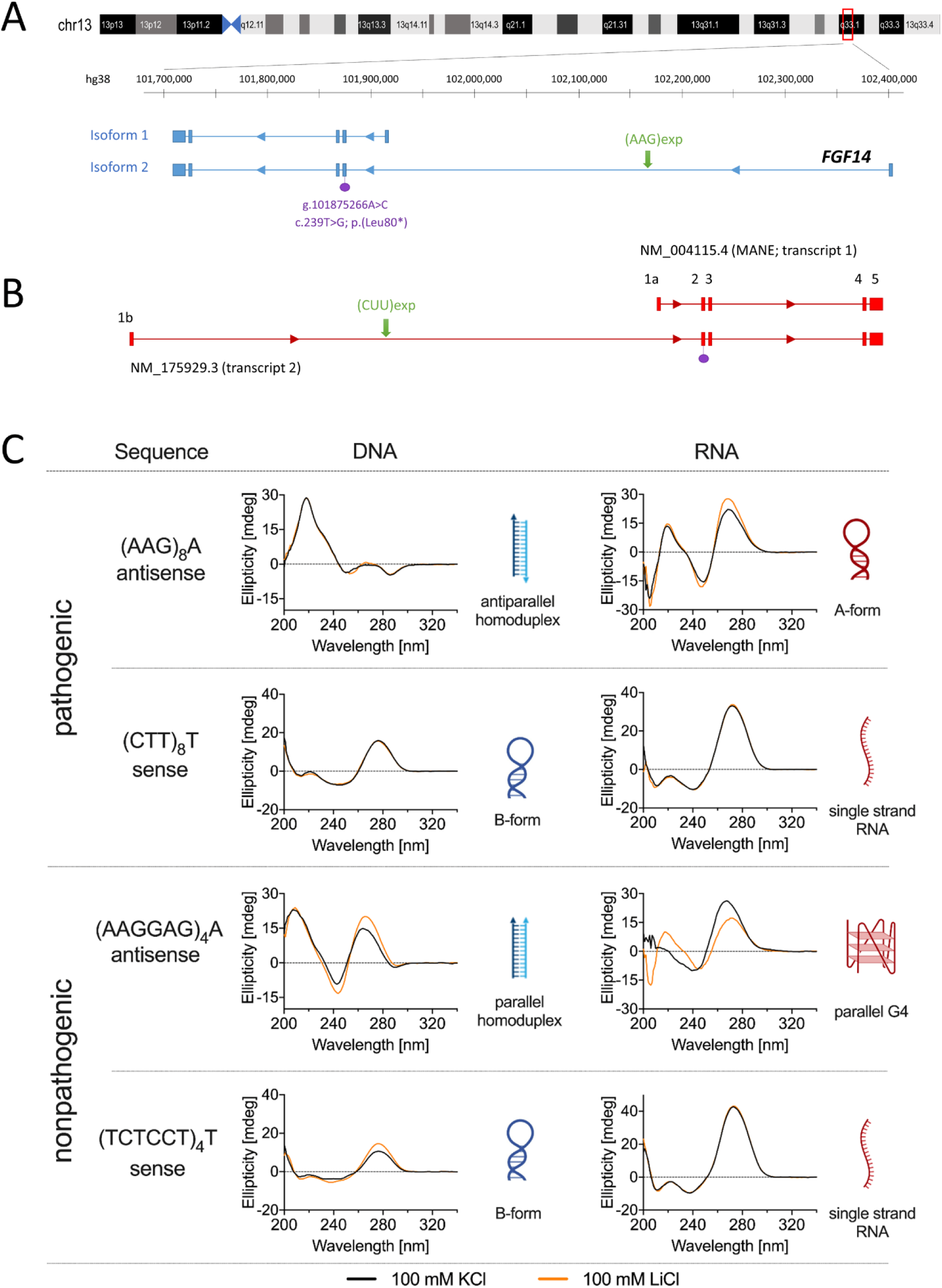
AAG and AAGGAG form different secondary structures at the DNA and RNA level. **A)** Schematic representation of the region on chromosome 13q33.1 containing the *FGF14* gene showing isoforms 1 and 2, which have alternative first exons. The gene is on the reverse strand. The green arrows show the location of the AAG expansion in intron 1 of isoform 2. The location of the novel nonsense variant (NM_175929.3: c.239T>G; p.Leu80*) reported in this study is indicated in purple. **B)** Schematic representation of *FGF14* pre-mRNA isoforms 1 and 2. The expansion (green arrow) is composed of CUU repeats in RNA context. **C)** Secondary structures formed by AAG and AAGGAG repeats at the DNA and RNA level, assessed by circular dichroism spectroscopy. AAG repeats form an antiparallel homoduplex whereas AAGGAG repeats form a parallel homoduplex at the DNA level. At the RNA level, the AAGGAG repeats fold into a parallel guanine-quadruplex (G4) while AAG repeats adopt an A-form RNA structure. On the contrary, the CTT and TCTCCT repeats adopt a B-form and CUU and UCUCCU repeats did not form any particular secondary structure under the tested conditions.

## Discussion

In this study, we identify intronic *FGF14* AAG expansions (SCA27B) as a major genetic contributor to cerebellar ataxia that had previously been overlooked by short-read technologies. Bioinformatics tools such as STRling^13^ can be used to detect this expansion from short-read genome data with a good predictive value (70%) and excellent sensitivity (100%; Supplementary Fig. 2). Although STRling largely underestimates the repeat count, the identification of a significant outlier value is an indicator of a possible expansion, requiring validation through an alternative method such as LR-PCR, RP-PCR and/or targeted long-read sequencing. A tool that can easily be implemented in routine diagnosis is ExpansionHunter^14^ using the STRipy interface^15^. When used with off-target/extended mode, this tool provides a closer estimation of repeat numbers but the values cannot be fully trusted and need validation by a more reliable method.

The frequency of pathogenic expansions found in our ataxia cohort, considering the previously established threshold of 250 repeats, was remarkably high (23%), equivalent to or even higher than previous reports^6,7,19–21,25,34–36^. The difference in incidence is possibly linked to the population studied but it could also result from the sensitivity of the genetic test used. We improved the LR-PCR conditions to minimize the amplification bias towards the smaller allele usually observed under standard conditions. This method could detect the largest expansion (937 repeats) reported so far. Current diagnostic strategies for SCA27B rely on sequential genetic tests based on the detection of normal and intermediate alleles (1000 bp being the usual limit of detection using fragment size analysis) combined with RP-PCR^37^. However, due to their inherent limitations, both tests may miss expansions that are unusually large and/or composed of other repeat motifs. A complete sequencing of *FGF14* alleles should thus be part of routine diagnostic procedures. Targeted nanopore sequencing is a cost-effective method that can easily be implemented to sequence *FGF14* expansions. Its known drawback is the higher error rate compared to other sequencing methods. However, errors can usually be easily distinguished from true sequence variations if the sequencing depth is sufficient, based on their random occurrence in single reads (errors) or presence in multiple reads (true variants). For example, nanopore sequencing leads to systematic errors (AAC instead of AAG) at the end of long reads. Moreover, smaller DNA fragments are preferentially sequenced by this technology. Since patients with *FGF14* repeat expansions show a high somatic variability that is correlated with expansion size, preferential sequencing of small fragments leads to underestimate repeat number in patients with large repeat expansions. To circumvent this problem, we used the median number of repeats instead of the mean, which is also closer to the size observed on gel.

The repeat expansion was mainly identified in patients with adult-onset cerebellar ataxia and was associated in 76% with a very recognizable phenotype that consisted of slowly progressive cerebellar ataxia with accompanying episodic symptoms of ataxia, often the initial manifestation, and/or downbeat nystagmus^38^. We found that episodic symptoms are not restricted to ataxia, but can also include episodic choreatiform movements, as described in *Fgf14* knock-out mice^39^. Likewise, mild pyramidal tract signs were present in one patient. In three cases, cerebellar ataxia was more severe. In one patient, an anxiety disorder likely contributed. In the two other cases, there was also marked cerebellar atrophy, usually not seen in SCA27B. The reason for this unusual severe atrophy remains unknown. It may indicate a second cerebellar disease, possible due to another genetic or non-genetic cause, or even a second hit that could be limited to the cerebellum. Two additional patients showed in later stages of the disease Parkinsonism and dementia, and dementia, respectively. In these two cases, another age-related neurological disease, idiopathic Parkinson’s disease in one and (likely vascular) encephalopathy in the other, were suspected. Of note, the patient with later onset of Parkinsonism and dementia previously had tentative clinical diagnosis of multiple system atrophy, cerebellar subtype (MSA-C). This would be the third patient with this diagnosis in whom a *FGF14* pathogenic expansion is identified^19,21^. However, in the context of our patient, a careful retrospective examination ruled out the MSA-C diagnosis and revealed that this patient in fact had a typical SCA27B phenotype with later onset of second diagnoses that are frequent in elderly people. Secondary disease as a possible confounder for SCA27B with fast progression has already been discussed^19^.

The mean AAO in our cohort was around 52.3 years, but it strikingly varied from 21 to 76 years. Although most patients have a late-onset of first symptoms, seven patients presented with an earlier form of the disease, with an onset before the age of 40. We observed a positive correlation between the number of AAG repeats and AAO, but the contribution of repeat number to AAO is rather limited. Accordingly, we observed an important individual variation independent of the number of AAG repeats, as illustrated by the two patients with 258 and 259 repeats, but started showing symptoms in early (30-35 years) and late adulthood (70-75 years).

The family history of patients with SCA27B implied incomplete penetrance, both in patients with 250-299 repeats and patients with ≥300 repeats. Incomplete penetrance above 300 repeats is further supported by the identification of pure repeat expansions above this threshold in two individuals of the HNR control population. Overall, the frequency of pathogenic *FGF14* expansions in the control population we studied, which is representative of the population from North Rhine-Westphalia, using 250 repeats as a threshold, is 7 out of 802 (0.87%). In particular, we observed two control individuals (46-50 and 66-70 years old) with a pure AAG repeat expansion ≥300 (313 and 319 repeats, respectively). We did not have details about their neurological status and these individuals may develop symptoms of SCA27B later in life. But it also suggests that incomplete penetrance exists within this range of repeats and that the threshold for complete penetrance, if it exists, may thus be higher i.e. 320-335 repeats, as suggested by Rafehi *et al*.^7^ Pellerin *et al*. demonstrated that *FGF14* repeat expansions tend to contract when inherited from the father, whereas they typically elongate when transmitted from the mother^6^. This aspect may at least partly explain the incomplete penetrance observed in some families and should be taken into account for genetic counseling.

We observed one family with a typical SCA27B phenotype in which the proband had 258 repeats and the affected parent 224 repeats. Genome sequencing did not reveal any other possible cause for the ataxia in this family. Furthermore, we also observed a significant enrichment of intermediate alleles (180-249 repeats) in patients with ataxia compared to the control population. This strongly suggests that expansions below 250 may predispose to SCA27B, as also already suggested by Pellerin *et al*.^40^ and Hengel *et al*.^25^ Among the eight families with intermediate alleles, two patients exhibited episodic symptoms and a possible SCA27B phenotype. However, the phenotypes appeared more heterogeneous, suggesting that some patients either had an additional or a different genetic cause. Additional causes include a pathogenic SCA6 expansion. Intermediate alleles could thus be contributing to the disease only in association with other factors. However, a specific analysis including a large number of such patients is needed to confirm or rule out the pathogenicity of intermediate alleles. Altogether, our observations currently support a reassessment of pathogenic thresholds to 220 for pathogenic alleles with incomplete penetrance and at least 320 for full penetrance. Genetic modifiers influencing the AAO and penetrance are likely multiple and possibly involve genetic variations at genes encoding DNA repair genes, like in other repeat expansion disorders^41^. Nevertheless, the observation of several cases with biallelic expansions also suggests that the size of the smaller allele and/or other eQTLs at the locus play an additive effect and partially determine the penetrance of alleles <320 repeats.

The pathophysiological mechanism associated with *FGF14* expansions is likely a loss-of-function of isoform 2. This is supported by similar clinical features in patients with point mutations and repeat expansions. The p.Phe150Ser (F150S; F145S in isoform 1) variant, which was the first variant identified in *FGF14,*^3^ also leads to the loss of interaction with voltage-gated channels and a loss of FGF14 capacity to control their activity^42^. However, patients with nonsense or the F150S variant are on average earlier affected, suggesting that the loss-of-function of isoform 2 may be incomplete as a result of the expansion (i.e. expanded alleles would be hypomorphic compared to point mutations). Another possibility is that loss-of-function of isoform 2 would occur only when the repeats extend beyond an even higher threshold in neurons, as a result of aging. This hypothesis could explain the clinical variability observed in patients and also that intermediate alleles have a probability to expand beyond the pathogenic threshold only in patients who have an overall lower control of microsatellite stability. It is also possible that the number of repeats detected in peripheral tissues such as blood might not always reflect the number of repeats present in the brain or cerebellum.

The somatic variability of pure AAG repeat expansions is remarkable. Although positively correlated with expansion size, we also see an individual variability of this phenomenon. In particular, somatic variability is influenced by the presence of interruptions and by the sequence of the 5’ flanking region (Supplementary Fig. 4D; Fig. 4B). The association of specific flanking regions with repeat stability or instability has already been reported by Pellerin *et al*.^43^ In this study, we extend this observation by showing that the four cytosines present in the flanking region associated with small alleles are crucial in this process. We hypothesize that these cytosines control the overall ability of the repeats to form secondary structures favoring repeat number amplification. This observation is to relate to secondary structures possibly formed by the repeats. We detected differences in the ability of pure pathogenic AAG and non-pathogenic AAGGAG repeats to form secondary structures at the DNA level but also at the RNA level. Noteworthy, the non-pathogenic AAGGAG RNA sequences can form a parallel G4 while pathogenic AAG repeats form an A-form RNA. G4 structures formed by other repeat expansions, including those found in *RFC1*/CANVAS, have been previously linked to a decrease in gene expression^44^. Furthermore, intronic AAG repeat expansions leading to Friedreich ataxia were shown to form R-loops that impair the transcription of *FXN* ^45–47^. However, AAG is the repeat motif present in *FXN* whereas in *FGF14*, the repeats present at the RNA level are CUU repeats (Fig. 7B). Hence, in the context of *FGF14*/SCA27B, there is a potential scenario where the capacity of AAGGAG repeats to generate G4 structures serves as a protective mechanism against the formation of other structures, such as R-loops on the complementary strand. A more comprehensive investigation of how these secondary structures affect gene expression is essential for *FGF14*, but requires studying this effect in the appropriate tissue or cell type, given the predominant expression of this gene in the brain and cerebellum.

In conclusion, this study reveals the complete sequence of pathogenic and non-pathogenic *FGF14* expansions. We suggest that pure AAG expansions are pathogenic from a lower threshold (comprised between 180 and 220 repeats) and account for at least 23% of patients with adult-onset cerebellar ataxia in European populations while interrupted expansions or expansions composed of other hexanucleotide repeats motifs are non-pathogenic. For this reason, diagnostic tests should incorporate not only the assessment of repeat numbers but also include a comprehensive sequencing of the expansion.

## Resources

Ensembl: https://www.ensembl.org

Uniprot: https://www.uniprot.org/

GTEx: https://www.gtexportal.org/

STRipy: https://stripy.org/

## Data availability

All data and scripts included in this study, with the exception of genome data, are available from the Supplementary tables or in the Github repository. The consent forms of the EXPAND study do not allow depositing genome data in a public repository. Pedigrees, age at onset, sex and family relationships have been removed from the manuscript as they were considered by MedRxiv as possible identifiable information but are available upon reasonable request.

## Data Availability

All data and scripts included in this study, with the exception of genome data, are available from the Supplementary tables or in the Github repository.

https://github.com/kilpert/FGF14_basecalling.git

https://github.com/kilpert/FGF14_analyses.git

## Acknowledgements

We thank all patients, family members, and control subjects who participated in this study. We also thank Nele Bohne and Dr. Theresa Kühnel for their contributions to establishing the LR-PCR analysis and Dr. Christine Beuck for her expert technical guidance with the CD spectroscopy measurements.

