## Supplementary material for "Advancing molecular, phenotypic and mechanistic insights of *FGF14* pathogenic expansions (SCA27B)"

### Supplementary Figures

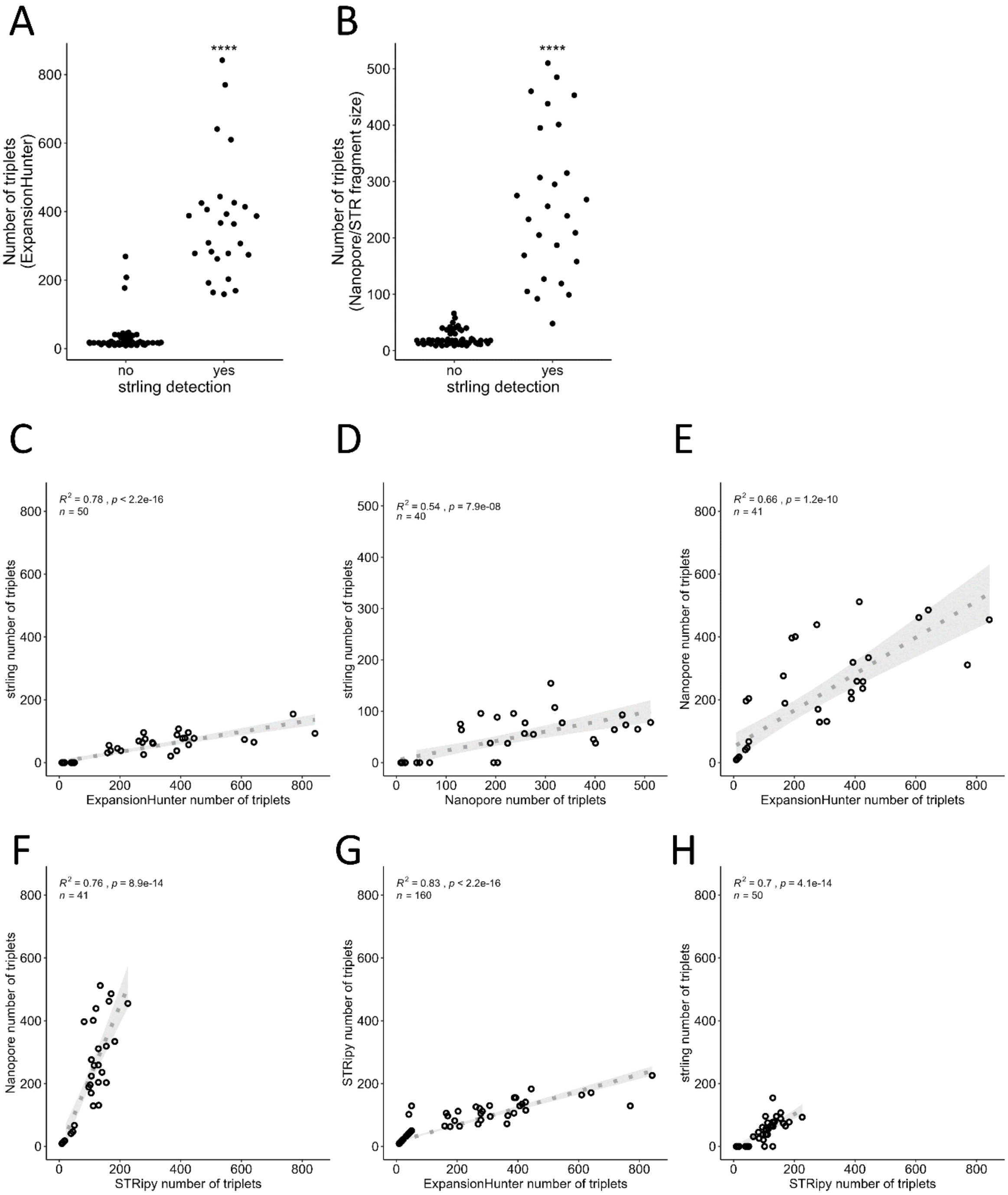


**Supplementary Figure 1**. **Comparison of the ability of bioinformatics tools to detect *FGF14* repeat expansion and repeat numbers**. **A)** Number of triplets detected by ExpansionHunter in samples for which a significant outlier value was or not detected by STRling. **B)** Median number of repeats detected by nanopore sequencing in samples for which a significant outlier value was detected or not by STRling. **C)** Correlation between the number of repeats detected by STRling and the repeat number estimated by ExpansionHunter. **D)** Correlation between the median number of repeats detected by nanopore sequencing and repeat number estimated by STRling. **E)** Correlation between the median number of repeats detected by nanopore sequencing and repeat numbers estimated by ExpansionHunter. **F)** Correlation between the median number of repeats detected by nanopore sequencing and repeat numbers estimated by STRipy. **G)** Correlation between the median number of repeats detected by STRipy and repeat number estimated by ExpansionHunter. **H)** Correlation between the median number of repeats detected by STRipy and repeat numbers estimated by STRling.

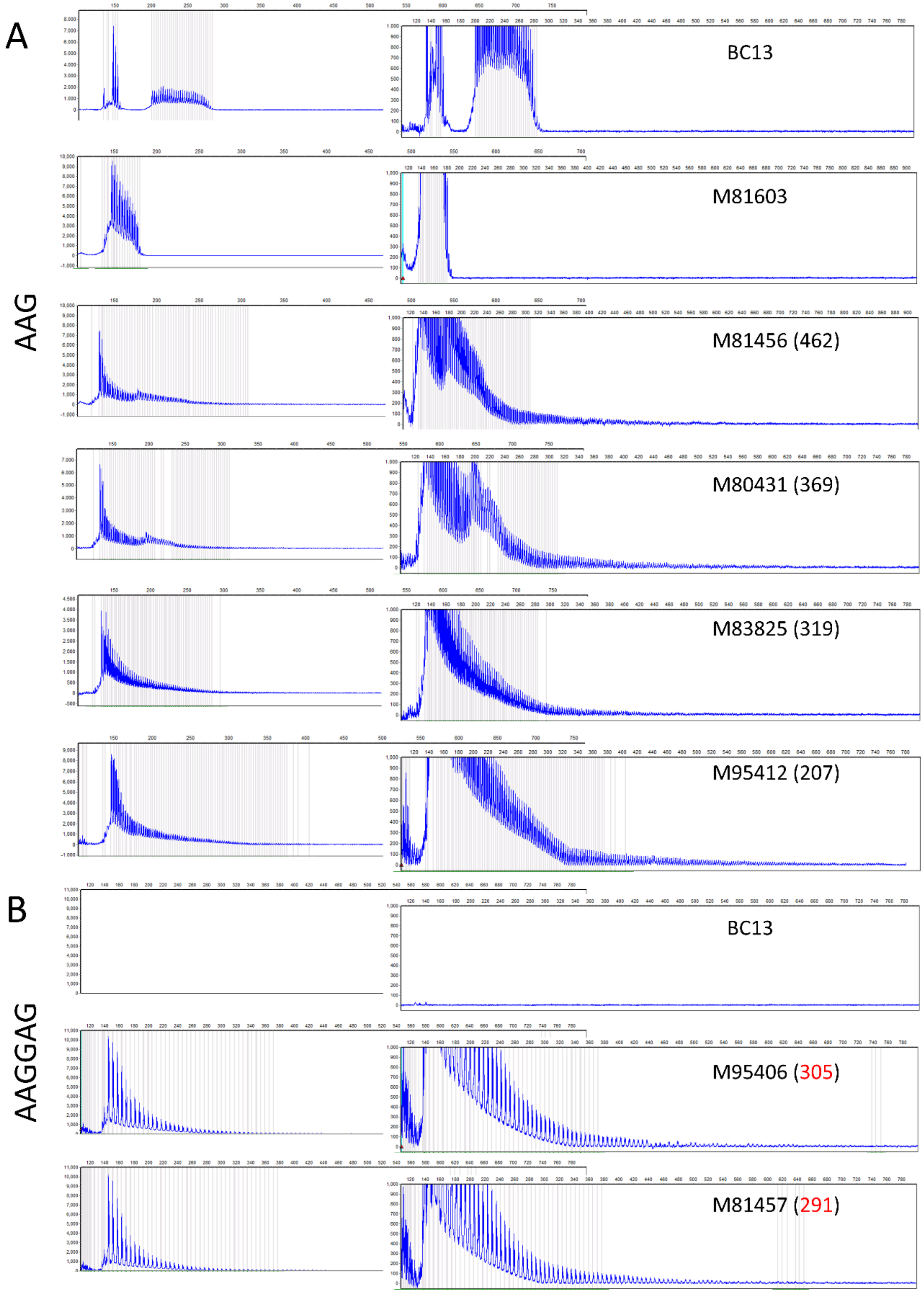


**Supplementary Figure 2. Limited ability of repeat-primed PCR (RP-PCR) to distinguish intermediate from pathogenic alleles. A)** RP-PCR profiles using AAG primer for a control individual (BC13) and five patients with cerebellar ataxia. M81603 is negative. The other four patients show a positive profile that cannot be used to distinguish with precision patients with alleles below the pathogenic threshold, intermediate alleles, and pathogenic repeat expansions. **B)** RP-PCR profiles using AAGGAG primer for a control individual (BC13) and two patients with cerebellar ataxia with an AAGGAG expansion. Patient IDs are not known to anyone outside the research group.

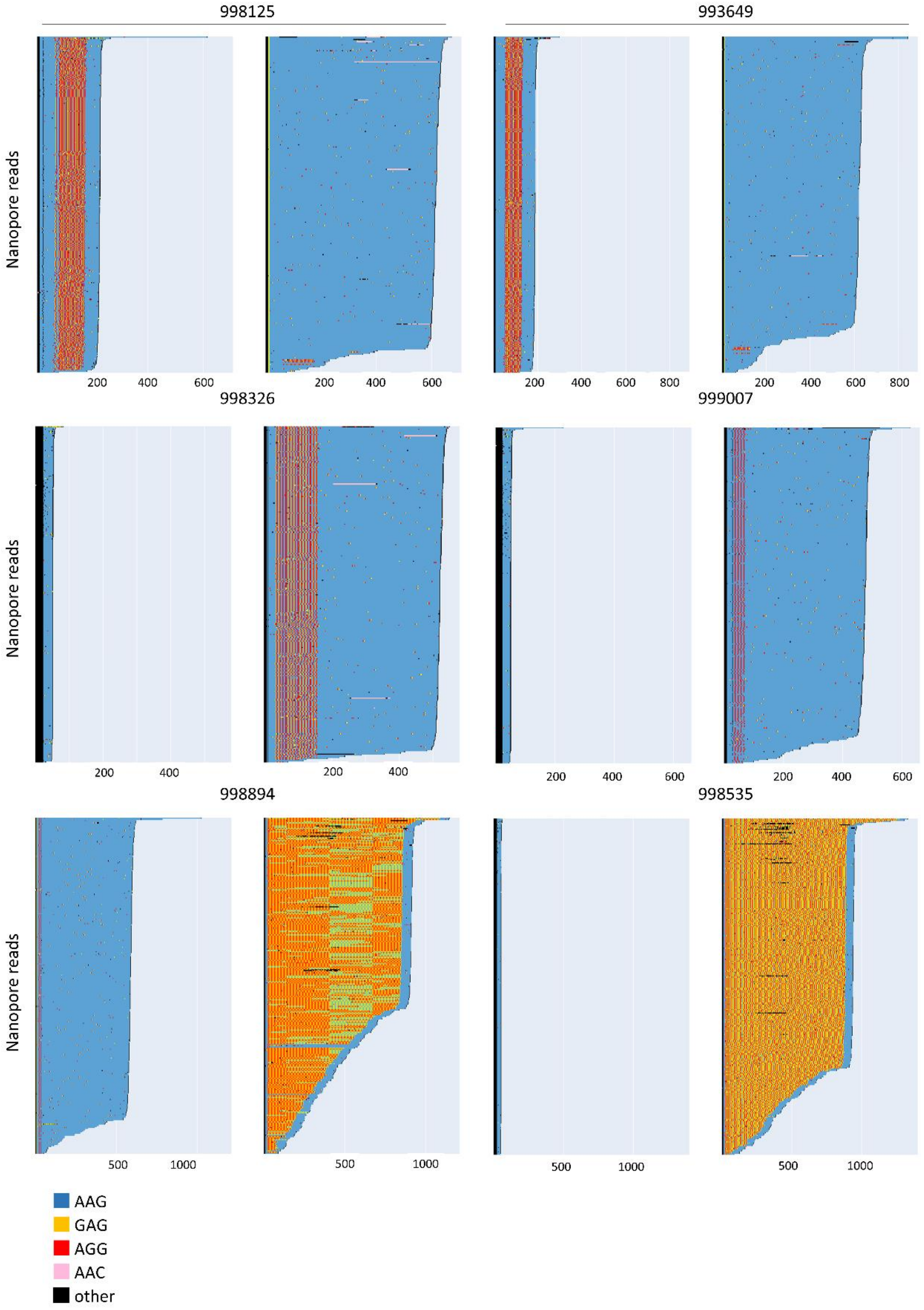

998125
993649
998326
999007
998894
998535
Nanopore reads
Nanopore reads
Nanopore reads
200 400 600
200 400 600
200 400 600 800
200 400 600 800
200 400
200 400
200 400 600
200 400 600
500 1000
500 1000
500 1000
500 1000
AAG
GAG
AGG
AAC
other

**Supplementary Figure 3. Examples of nanopore read profiles detected in control subjects.** An increase frequency of interrupted alleles or alleles with AAGGAG expansions was detected in the control populations compared with patients with ataxia.

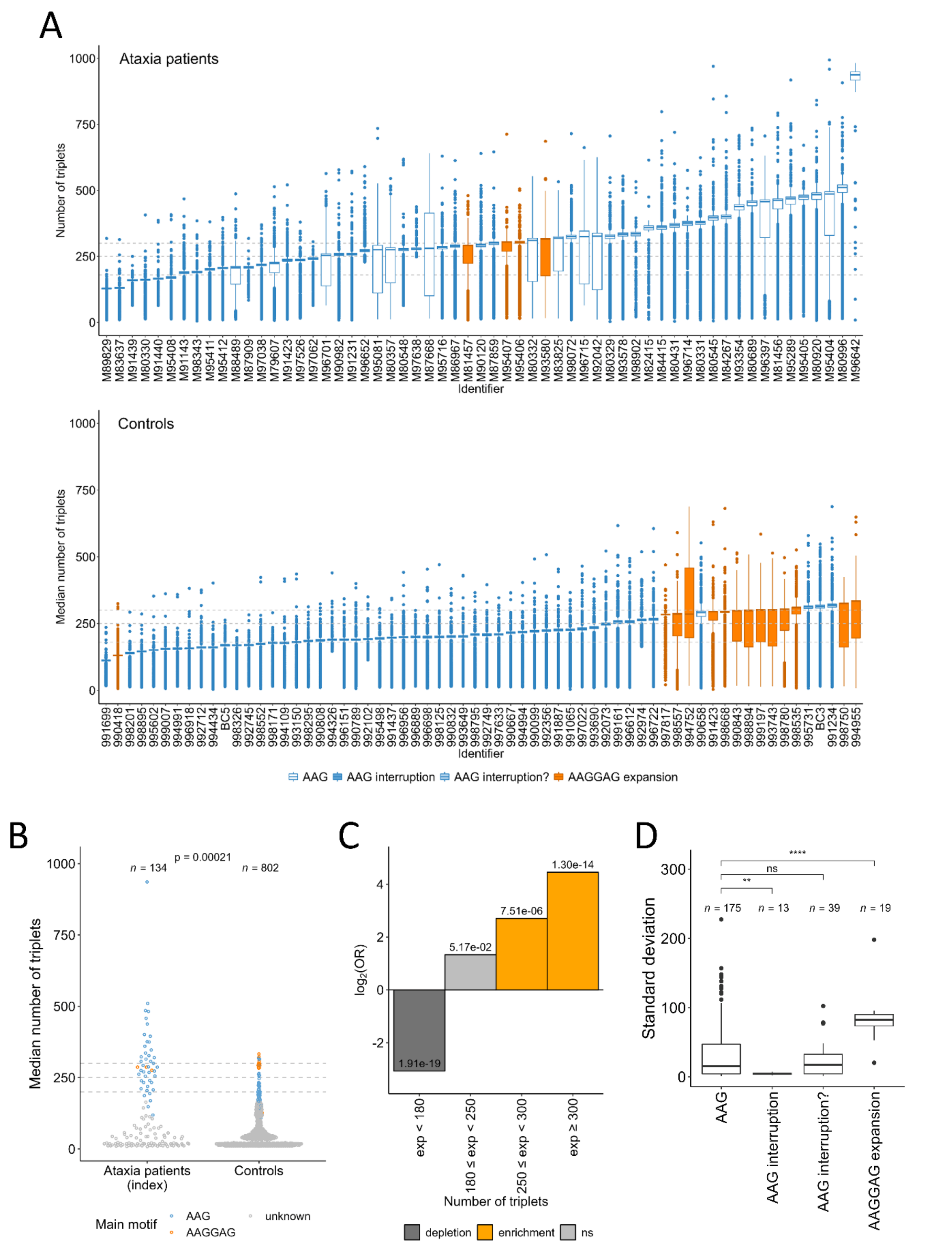


**Supplementary Figure 4**. **Distribution of *FGF14* alleles in patients with cerebellar ataxia and control subjects. A)** Box plots showing the distribution of the number of triplets on both alleles calculated from nanopore data for the 59 patients with ataxia and 64 control individuals sequenced by nanopore sequencing. Pure AAG alleles are depicted in blue; AAGGAG alleles in orange. Patient IDs are not known to anyone outside the research group. **B)** Comparison of the median sizes of the larger allele in patients with cerebellar ataxia and control subjects. **C)** Log odds ratio according to repeat numbers (ataxia patients versus controls) showing a significant enrichment of larger alleles > 180 repeats in patients with cerebellar ataxia. **D)** Standard deviation as a measure of somatic variability for alleles with pure AAG repeats, alleles with interruptions limited to 3’ or 5’, true interruptions (disrupting repeats), and alleles with AAGGAG repeats.

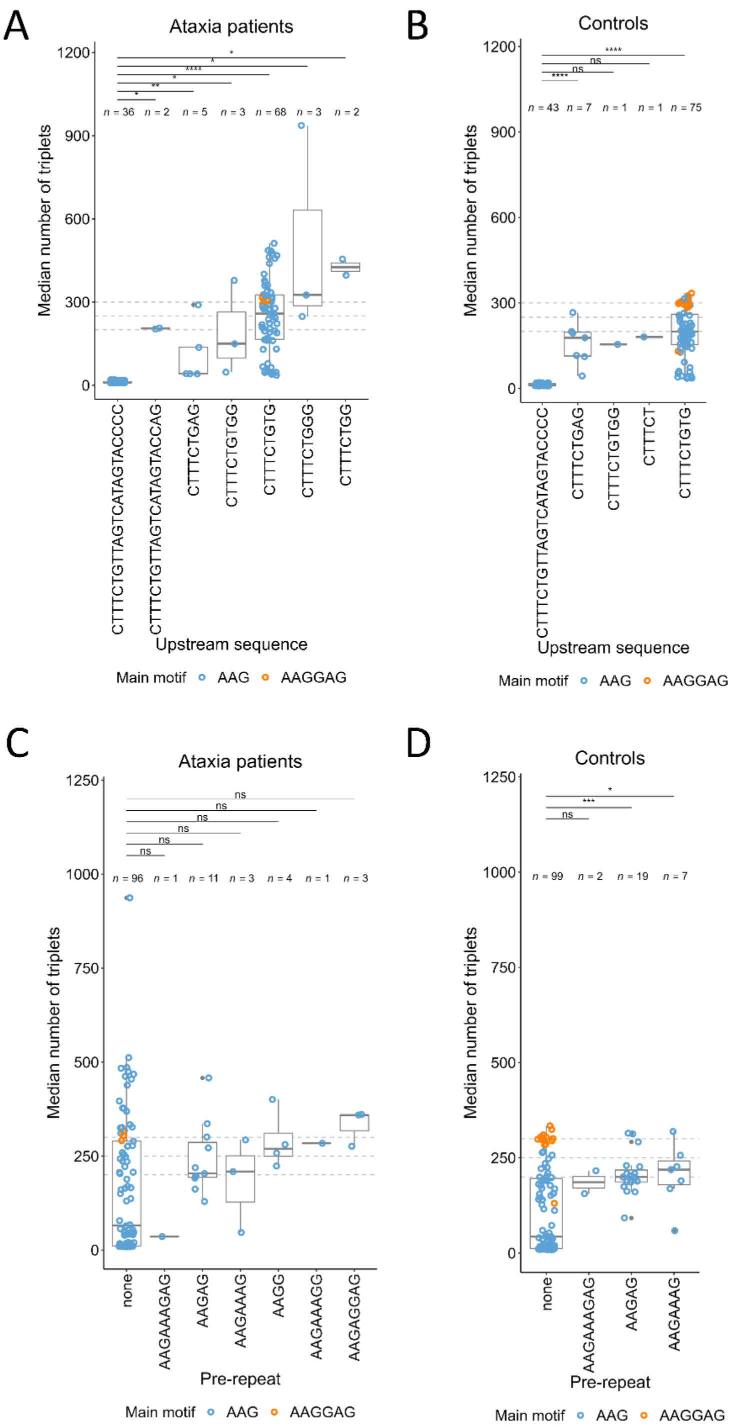


**Supplementary Figure 5**. **Effect of 5' flanking regions on repeat instability**. **A)** and **B)** Median number of triplets for each allele according to the flanking region sequence in patients with ataxia (A) and in control subjects (B). **C)** and **D)** Median number of triplets for each allele according to the pre-repeat motif in patients with ataxia (C) and in control subjects (D).

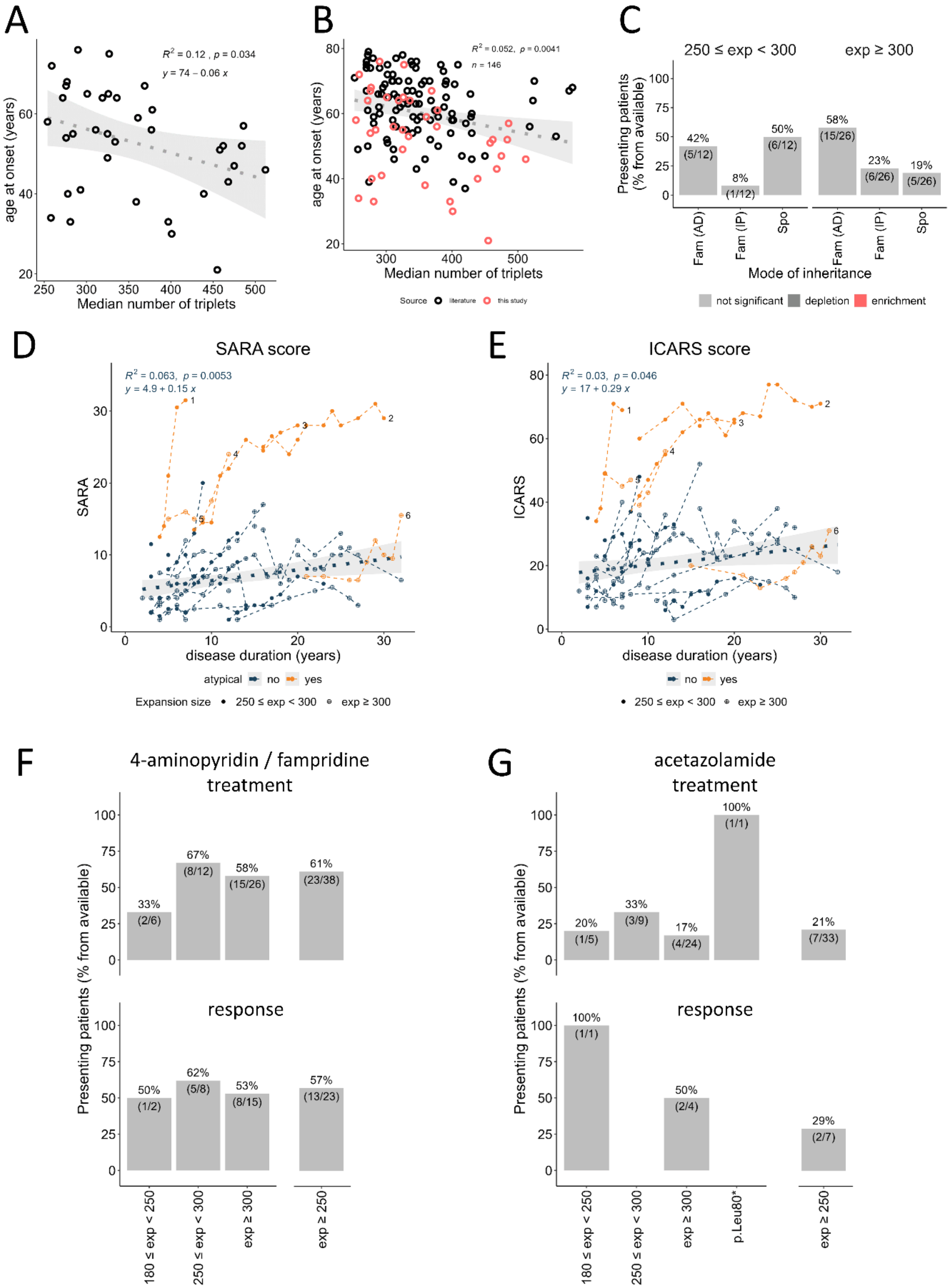


**Supplementary Figure 6**. **Additional clinical comparisons**. **A)** Correlation between the age at onset and *FGF14* AAG repeat number including only patients from this study, excluding outlier.

**B)** Correlation between the age at onset and *FGF14* AAG repeat number taking all patients from this study (red) and patients from previous studies (black) into account, excluding outlier. **C)** Comparison of family history presentation according to expansion size. **D)** SARA scores of patients with *FGF14* repeat expansions. **E)** ICARS scores of patients with *FGF14* repeat expansions. In both graphs shown in panels E) and F), atypical patients appear in orange while the other appear in blue. Scores from the same patients at different time points are connected with dashed lines. Numbered last data points mark lines corresponding to atypical patients. **F)** Bar graphs showing the response to 4-aminopyrimidine/fampridine treatment in patients with intermediate alleles, patients with 250 to 299 repeats and patients with ≥ 300 repeats (left). On the right, the graph shows the response for all patients ≥ 250 repeats. **G)** Bar graphs showing the response to acetazolamide treatment in patients with intermediate alleles, patients with 250 to 299 repeats, and patients with ≥ 300 repeats (left). On the right, the graph shows the response for all patients ≥ 250 repeats.

### Supplementary methods

***Genome sequencing.*** Libraries were prepared with the DNA tagmentation based library preparation kit (Illumina) without PCR, with 300-500 ng genomic DNA input. Library preparation was followed by clean up and/or size selection using SPRI beads (Beckman Coulter Genomics). After library quantification (Qubit, Life Technologies) equimolar amounts of library were pooled. The library pools were quantified using the Peqlab KAPA Library Quantification Kit and the Applied Biosystems 7900HT Sequence Detection System and then sequenced on an Illumina NovaSeq6000 sequencing instrument with a paired-end 2x150bp protocol. Raw sequencing data underwent preprocessing using cutadapt[48] to remove adapter sequences. Read mapping used bwa-mem[49], bwa-mem 2[50], and bwa-meme[51]. Duplicate reads were removed with samblaster[52]. Sorted and indexed CRAM files were generated by samtools[53]. In an initial phase, we compared the ability of ExpansionHunter DeNovo[54] and STRling[33] to detect known repeat expansions from short-read genome data including TTTTA/TTTCA repeat expansions in *MARCHF6*[55] and *STARD7*[56], and a full (>200) CGG repeat expansion in *FMR1* (Fragile X). Both tools performed similarly but we chose STRling based on its ability to detect a more accurate number of repeats compared to ExpansionHunter DeNovo. We then used STRling (version 0.5.2) to call short tandem repeats on the processed and mapped sequencing data at the genome-wide level. Known repeat expansions were additionally called by ExpansionHunter[24] using the repeat expansion JSON file downloaded from STRipy[14] using the extended mode (i.e. with off-target regions).

***Long-range PCR amplification***. Repeat expansions at the *FGF14* locus were amplified by Long-Range PCR (LR-PCR) from genomic DNA extracted from blood using a protocol adapted from Rafehi *et al*[8]. The amplification of *FGF14* repeat alleles was performed from 50 ng genomic DNA in 25 µl using the HotStarTaq DNA Polymerase (Qiagen, Hilden, Germany) and 0.20 µM of each of the following primers: FGF14_RPP_ F1: AGCAATCGTCAGTCAGTGTAAGC; FGF14_LRP_ R1: CAGTTCCTGCCCACATAGAGC. The PCR program comprised an initial step at 95°C of 15 min; followed by 28 cycles, each consisting of 30 seconds at 95°C, 30 seconds at 60°C and 2 minutes at 72°C; and a final step at 72°C of 10 min. LR-PCR amplicons were analyzed on a 1.3% agarose gel. The PCR was performed with a FAM-marked forward primer for gene fragment analysis on ABI 3130xl DNA Analyzer (Applied Biosystems, Waltham, MA). The size of FGF14 alleles below 700-1200 bp were quantified using the GeneMarker software (SoftGenetics LLC, PA).

***Targeted sequencing with Oxford Nanopore Technology (ONT)***. Nanopore sequencing of LR-PCR products was performed for subjects showing an allele above 700 bp. LR-PCR amplification was performed using the same protocol (without FAM marked primer) in a total volume of 75 µl. LR-PCR amplicons were purified using the DNA Clean & Concentrator (Zymo Research). We use the SQK-LSK109 ligation-based sequencing kit (Oxford Nanopore) and the native barcoding protocol (EXP-NBD196) to prepare the libraries and multiplex samples, respectively. In brief, this procedure involves the following stages: 1) End-prep, where 200 fmol of each purified amplicon undergoes incubation with NEBNext Ultra II End Repair/dA-tailing Module Reagents at 20°C for 5 min and 65°C for 5 min in a 96-well plate; 2) Native barcoding ligation, comprising incubation with native barcodes and NEB Blunt/TA Ligase Master Mix at 20°C for 20 min and 65°C for 10 min; 3) Pooling of barcoded amplicons and purification using AMPure XP beads (Beckman Coulter); 4) Adapter ligation, involving a 10-minute incubation with Adapter Mix II Expansion/NEBNext Quick Ligation Reaction Module followed by clean-up with AMPure XP beads. All steps were executed in accordance with the manufacturer's recommendations. Approximately 15 ng of the final prepared library was loaded onto a MinION Mk1B R9.4.1 FLO-MIN106 flow cell, and nanopore sequencing was conducted for up to 24 hours, and monitored using the MinKNOW software.

Basecalling and analysis of nanopore data were performed using command line Snakemake[15] workflows available at GitHub (https://github.com/kilpert/FGF14_basecalling.git; https://github.com/kilpert/FGF14 _analyses.git). This workflow takes fast5 files as input and utilizes several tools to generate sequence_summary.txt, final_summary.txt, and fastq.gz files. Guppy[57] (version 6.4.6) was used for basecalling, pycoQC[58] (version 2.5.2) and NanoPlot[59] (version 1.41.6) for quality control. The command line version of Guppy, guppy_basecaller, used the parameters: "“`--recursive --compress_fastq --do_read_splitting --calib_detect --records_per_fastq 0 --enable_trim_barcodes`". After comparing the basecalling hac (dna_r9.4.1_450bps_hac.cfg) and sup (dna_r9.4.1_450bps_sup.cfg) models, the "sup" model was selected for further analysis. The "pass"-reads of samples sequenced on multiple runs were pooled.

The initial fastq files were quality trimmed and filtered with BBMap[60] bbduk.sh using the parameters: "`-Xmx2g qin=33 minlen=200 qtrim=lr trimq=10 maq=10 maxlen=100000`". Two 25 bp flanking sequences upstream chr13(hg38):102161532-102161557, ATATCAATATTCTCTATGCAACCAA) and downstream (chr13:102161726-102161751, TAGAAATGTGTTTAAGAATTCCTCA) of the repeat expansion were used to filter for all reads that had both flanking sequences, allowing 2 mismatches per flanking sequence using bbduk.sh with --literal and --edist parameters. Reads where both flanking sequences showed a -strand orientation were subsequently converted to their reverse-complement (+strand) sequence. All other reads were discarded. In the next step, flanking sequences were trimmed off from both sides using Cutadapt[48] to leave only the repeat containing region, as well as the invariable and variable region (5' flanking region) and pre-repeat. We considered only reads containing 2x AAG (i.e. AAGAAG). The workflow calculates statistics and generates plots (custom Python and R scripts) to characterize the nature of the repeat expansion in length and motif composition. Specific alleles (length and motif) can be defined manually for enhanced visualization.

For each sample, reads were visually inspected in Geneious Prime® 2019 (Biomatters Ltd.) for identification of the sequences corresponding to the invariable region, 5' flanking region, pre-repeat, main motif, additional repeat motif, and interruptions. Differences between alleles (inclusion or exclusion of sequences, minimum or maximum length) were used to separate reads from different alleles into a1 (smaller), a2 (larger) and a3 (intermediate, mosaic cases). Plots were generated for the separated alleles using up to 300 random reads per allele. For each allele and sample, we calculated the median size of the repeat region (after subtracting from the length of each sequence, the sizes of the invariable region, 5' flanking region and pre-repeat), the median number of triplets and its standard deviation. For comparisons with the sizes obtained by fragment analysis, we increased the median size by 146 bp, which were previously removed by trimming.

***Repeat-primed PCR.*** *FGF14* AAG repeat expansions were amplified by repeat-primed PCR (RP-PCR) with 6-FAM labeled FGF14_RPP_F1 (AGCAATCGTCAGTCAGTGTAAGC), and non-labeled RPP_M13R (CAGGAAACAGCTATGACC) and FGF14_RPP_AAG_RE_R1 (CAGGAAACAGCTATGACCCTTCTTCTT-CTTCTTCTTCTT). AAGGAG expanded alleles were amplified using FAM-FGF14_RPP_F1, P3 (TACGCATCCCAGTTTGAGACG), and

P3_AAGGAG (TACGCATCCCAGTTTGAGACGAAGGAGAAGGA-GAAGGAGAAG). PCR was performed from 100 ng genomic DNA, with 0.8 µM primer FGF14_RPP_ F1, 0.8 µM primer RPP_M13R or P3, and 0.26 µM primer FGF14_RPP_AAG_RE_R1 or P3_AAGGAG using the HotStarTaq Master Mix (QIAGEN). The PCR program consisted in 95 °C for 15 min, followed by 35 cycles (94 °C for 30 s, 61 °C for 30 s, and 72 °C from for 2 min) and a final extension step at 72 °C for 10 min. RP-PCR products were detected on an ABI 3130xl DNA Analyzer and analyzed using GeneMarker software (SoftGenetics).

***Statistics.*** Fisher's tests (two-sided) were performed to determine associations between: (i) a sample belonging to a class of triplet numbers (either in all alleles or just in the larger allele of an individual) and being an ataxia patient; (ii) a patient belonging to a class of *FGF14* pathogenic expansion size or point mutations and presenting certain symptoms; (iii) a patient belonging to a class of *FGF14* pathogenic expansion size and having a certain family history presentation; and (iv) a patient belonging to a class of *FGF14* pathogenic expansion size and having a certain response to treatment. Odds ratios were $\log_2$ transformed and indicate enrichment or depletion, for positive or negative values, respectively. *P*-values were adjusted for multiple comparisons using Bonferroni correction. Mann-Whitney U test (two-sided) followed by Bonferroni correction for multiple testing (when applicable) was used to assess (i) the median number of repeats difference between ataxia patients and controls; (ii) the age at onset / age at last examination / disease duration difference between various classes of *FGF14* pathogenic expansion size and point mutations; (iii) the difference in the number of triplets quantified by ExpansionHunter or Nanopore/STR fragment size for samples for which STRling detected or not FGF14 expansions; and (iv) the standard deviation of the number of triplets difference between various categories of main repeats and interruptions. The details and results of all statistical tests performed appear in Supplementary Table 2.

***Circular dichroism spectroscopy.*** The AAG and AAGGAG repeats and the complementary counterparts were purchased as 25-mer DNA and RNA oligos from Microsynth (Switzerland) and dissolved in nuclease-free water at 100 µM. The sequences of the oligos are displayed in Fig. 8C and are identical for DNA and RNA except that thymidines were replaced by uracils in RNA. The final concentration of oligos was 50 µM in 10 mM cacodylate buffer (pH 7.2) either supplemented with 100 mM $K^+$ or $Li^+$ as indicated. Secondary structure formation was carried out by heating the oligos to 95 °C for 5 min and slowly decreasing the temperature with a ramp rate of 0.01 °C/s to 20 °C using the LightCycler® LC480II (Roche). The folded oligos were kept at 4 °C overnight and measured the next day. Circular dichroism (CD) spectra were measured over a spectral range of 200-340 nm on a Jasco J-710 CD spectropolarimeter coupled to a Jasco PFD-3505 Peltier temperature controller. All measurements were carried out at 20 °C in a quartz cuvette with a 1 mm path length using a scanning speed of 200 nm/min, a response time of 2 s, a bandwidth of 1 nm, and an accumulation of four spectra.
